# Stepping in Place (SIP) as a Novel Physical Ability Assessment in Neurorehabilitation: A Wearable Device-based Validation Study

**DOI:** 10.1101/2024.04.30.24306653

**Authors:** Bin Hu, Doreen Amini, Izma Ghani, Abdul-Samad Ahmed, Shahryar Wasif, Taylor Chomiak

**Author notes:** Correspondence concerning this article should be addressed to Professor Bin Hu MD. Ph.D., Suter Professor for Parkinson’s Disease Research, Founder and Director, OpenDH program, Department of Clinical Neuroscience, University of Calgary, Alberta, Canada.

## Abstract

The assessment of physical ability is a critical component in developing personalized exercise prescriptions, monitoring disease progression, and evaluating intervention outcomes across various clinical and general populations. This study evaluates how objective physical performance parameters, measured during a stepping in place (SIP) exercise via Ambulosono wearable system, relate to subjective perceptions of fatigue and breathlessness using Borg and Fatigue Scores. Our overall results show that SIP, as a convenient and simple exercise modality, can be used to rank a user’s physical ability level based on both objective and subjective parameters. Furthermore, while the objective walking/gait parameters may have some predictive ability for the such parameter as cadence, they do not appear to significantly predict the subjective fatigue or breathlessness scores, either before or after the activity. This lack of significant relationships suggests that factors other than the measured objective gait metrics may play a more important role in determining subjective experiences of fatigue and breathlessness during the stepping exercise.

## Introduction

The assessment of physical ability is a critical component in developing personalized exercise prescriptions, monitoring disease progression, and evaluating intervention outcomes across various clinical and general populations (Rahman et al., 2019; Zheng et al., 2022). Traditional approaches and recent studies using fitness tracking devices have increasingly relied on objective measures of physical performance, such as walking distance, speed, and cadence, obtained through standardized tests or wearable sensor technologies (Smith et al., 2017; Brown et al., 2020). These objective metrics were often used to provide quantifiable indicators of functional capacity and fitness levels and to promote recommendations in public health. However, the relationship between these objective measures and subjective experiences like fatigue and breathlessness remains incompletely understood (Davis et al., 2015; Thompson & Bailey, 2022).

While some studies have reported associations between objective physical activity levels and subjective fatigue or breathlessness ratings (Davis et al., 2015), others have found weak or inconsistent correlations (Wilson et al., 2018). This discrepancy highlights the complex and multidimensional nature of physical ability assessment, encompassing not only objective performance outputs but also subjective physiological and psychological responses to exercise (Green et al., 2017; Liu et al., 2020).

Recent advances in wearable sensor systems, such as Ambulosono, have enabled more detailed and precise capture of gait parameters during activities like Stepping in Place (SIP) exercises (Hu et al., 2019). These technologies provide an opportunity to comprehensively evaluate objective measures of physical performance alongside subjective self-reported experiences. Understanding the interplay between these objective and subjective components is crucial for developing targeted exercise interventions tailored to individual needs and capabilities (Santos et al., 2021).

SIP emerges as a practical and accessible exercise modality that transcends the confines of traditional fitness paradigms (Chomiak et al., 2014, 2015, 2015, 2017, 2019). Seamlessly integrating into the fabric of daily routines, SIP offers a unique blend of convenience, safety, and adaptability, catering to the diverse needs of populations spanning the spectrum of age, ability, and lifestyle. By eliminating the necessity for specialized equipment or environments, SIP presents a low-barrier entry point into the realm of physical activity, empowering individuals to reclaim their well-being on their own terms (Hu et al., 2019).

Stepping exercises also represent a valuable model for investigating physical ability assessment, as they involve controlled, rhythmic movements that can be objectively quantified while also eliciting subjective responses related to fatigue and breathlessness. Moreover, instrumented stepping exercises facilitated by wearable gait analysis systems have shown promise for home-based rehabilitation in populations with conditions like Parkinson’s disease (Hu et al., 2019).

In this study, we conducted a comprehensive analysis of objective gait parameters measured by the Ambulosono system during a stepping exercise, and examined their relationship with subjective fatigue and breathlessness experiences reported by participants. Our data analytics included two complementary approaches to categorization or grouping of the physical abilities among participants: parametric similarities of data variables (e.g. via clustering, principal components and machine learning techniques) and predictive statistics (e.g. via correlation and regression analyses). Specifically, our goals were to:

- Validate the structure and underlying dimensions of the objective gait performance measures obtained from the wearable sensors (Chen et al., 2021; Wang et al., 2023).
- Identify distinct clusters of participants based on their objective physical performance levels during the stepping exercise (Brown et al., 2020; Khan et al., 2019).
- Assess subjective changes in fatigue and breathlessness before and after the exercise, and cluster participants based on their subjective response patterns (Khan et al., 2019).
- Determine the associations between the objective gait metrics captured by Ambulosono’s system and participants’ subjective experiences of fatigue and breathlessness (Thompson & Bailey, 2022; Garcia et al., 2021).

By integrating objective gait analysis from wearable sensors with subjective self-reported data, this study aimed to provide a nuanced understanding of physical ability assessment during a controlled stepping exercise. The findings have implications for developing multidimensional approaches that consider both objective performance outputs and subjective experiential components (Santos et al., 2021; Zheng et al., 2022). Such integrated assessments are essential for optimizing exercise prescription strategies, monitoring individual progress, and ultimately enhancing quality of life across diverse populations through personalized, holistic interventions (Rahman et al., 2019).

By establishing a robust and statistically significant relationship between these objective improvements and subjective experiences, our research aims to position SIP as an indispensable component of exercise prescriptions, public health initiatives, and holistic well-being strategies. Through the dissemination of our findings, we wish to advocate for the widespread adoption of SIP as a scientifically validated, sustainable, rewarding, and transformative form of physical activity that nurtures the body and mind (Chomiak et al., 2014, 2015, 2015, 2017).

## Methods

Our study was carried out via Open Digital Health (OpenDH) program, a student research training initiative aimed at providing early exposure to learning and applying advanced research and data analytics skills. Our overarching strategy centers on student education through the integration of advanced data analytics and statistical training, leveraging the capabilities of Generative Pre-trained Transformer (GPT) models.

The scalability of GPT-based training, such as ChatGPT-4, can significantly enhance accessibility to advanced statistics education. Students from diverse backgrounds and disciplines can access high-quality, interactive training resources online, breaking down barriers to advanced education in statistics and research. By providing an interactive, personalized learning environment, OpenDH program fosters a deeper understanding of statistical concepts among students, while simultaneously exposing them to cutting-edge AI tools and methodologies.

### Participants

The dataset included 24 undergraduate students, aged from 18-20 (mean=xxx) who were volunteers recruited from the University campus via OpenDH program. All subjects were free from injury and had no history of hospitalization or chronic disease influencing their exercise capacity. Moreover, they were not involved in any competitive sport and they were lifetime non-smokers. Ethics approval was obtained from the University of Calgary Research Ethics Board as part of Ambulosono registered trial (ISRCTN06023392). Informed written consent was obtained from participants at baseline prior to participation.

### Ambulosono Device and Data Capture

Ambulosono is a single sensor-based motion tracking device consisting of 3-axis MEMS-based gyroscopes and 3 accelerometers with validated accuracy (Chomiak et al. 2019). Ambulosono App uses fusion codes for automatic gravity calibrations and real-time angle output (pitch, roll, and yaw) with remarkable accuracy. This includes not only basic measures such as distance and time but also useful metrics like step height, cadence and step time. The device was attached to the single leg of a user, just above the patellar top line, through the use of a high-performance thigh band (Chomiak et al., 2019). The application utilizes sample sensor output at a rate of 50 Hz for step parameter calculations such as step length based on its ability to automatically record the angular velocity of hip flexion in real time and, after correction for baseline drifting, to calculate the step length (SL) of each step. All SIP data files are locally auto-saved, archived, and encrypted.

In this study, the step height (SH) during SIP is represented as a nominal value for both SL and proxy for the distance of forward locomotion. SH obeys a linear relationship with SL (Chomiak et al. 2015), i.e. SH equals 40.5 % of SL (see equations in ref. [Chomiak et al. 2015], where (SL = (Pd)(LL) and SH = (Pd)(LL)(0.405). The Pd represents the captured peak hip flexion and LL represents limb length as measured from the center of hip bone (iliac crest)to the floor (Chomiak et al., 2015).

### SIP Instructions

– Preparation: Before starting, participants receive a comprehensive briefing about the study objectives and the detailed exercise protocol. Wearable devices are provided to each participant to ensure accurate recording of physical performance metrics during the stepping exercise.
– Music Selection: Five playlists were created and presented to participants based on vocal signings by Canadian singers who are the inductees of Canadian Music Hall of Fame. They feature a wide range of music genres and tempos as well as cultural popularities such that their perceived motivational values differ significantly. During stepping if the music appeals to them, participants were encouraged to step more vigorously, increasing both the height and speed of their steps. This adjustment is intended to mirror a spontaneous increase in effort when engaging with preferred music.
– Subjective Assessment: Both before and after stepping to each song which was randomly presented from each playlist, participants were asked to report their levels of breathlessness and fatigue, using Borg Breathless Scales and Fatigue rating scores (Borg, 1998).

The analysis and findings concerning the influence of music on both objective and subjective measurements will be detailed in a separate paper describing music preferences on exercise engagement and the perceived exertion of participants.

## Data Analysis

Schematic of GPT Advanced Data Analysis Plan

**Figure 1.**
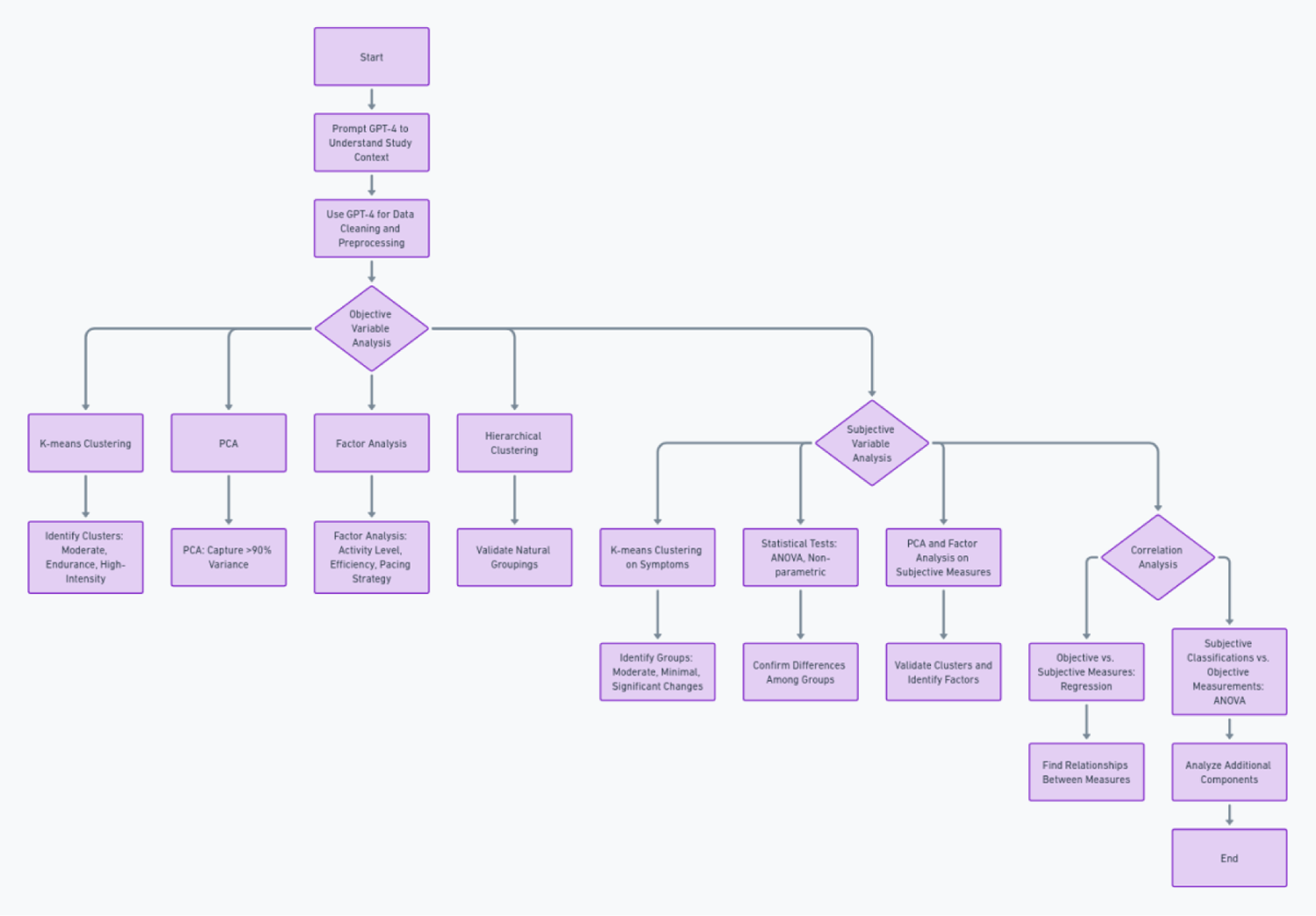
Schematic of statistical data analysis plan and its implementation via GPT-4 Advanced Data Analysis platform that supports automatic transformation of natural language instructions into python codes for statistical data analysis.

### Objective Variable Analysis

1. Clustering Analysis: - K-means clustering was used to identify three distinct clusters of data similarities based on objective performance metrics during the stepping exercise such as moderate performers, endurance-focused, and high-intensity performers.
2. Hierarchical Clustering was used to further support the natural groupings within the dataset, reinforcing the cluster validity.
3. Principal Component Analysis (PCA) and Factor Analysis (FA). PCS was used to capture the variances in the dataset after scaling and reveal the underlying dimensions related to overall activity level, exercise efficiency/intensity, and pacing strategy. Factor Analysis was used to further identify overall activity level, exercise efficiency/intensity, and the trade-off between efficiency and pacing strategy by further validating the underlying dimensions (factors) that explain the observed correlations among the variables. This dual approach helps in both dimensionality reduction and the identification of latent variables.

These methods are also widely used in clustering analysis to assess the appropriate number of clusters by considering within-cluster variance and the similarity of objects within their own cluster compared to other clusters, respectively.

### Subjective Variable Analysis

Clustering analysis and other methods mentioned above were also used to categorize participants into three distinct groups based on changes in fatigue and breathlessness scores that represent moderate increase, minimal changes, and significant increases in fatigue and breathlessness.

#### 2. Statistical Tests

ANOVA and non-parametric tests were used to confirm statistically significant differences among the clusters for fatigue and breathlessness changes. PCA and factor analysis was used to validate clusters in the reduced-dimensional space of PCA, and further identify the underlying factors explaining variations in subjective experiences.

### Objective and subjective variables correlation analysis

Regression analyses were used to reveal the relationships between objective walking parameters and subjective breathlessness scores before or after activities. Machine learning models were used to extract important features of the relationship. ANOVA and nonparametric tests were used to reveal the significant correlation and machine learning models between subjective and objective clusters and classifications and including the principal components from PCA.

## Results

### Descriptive Statistics

**Table 1.**
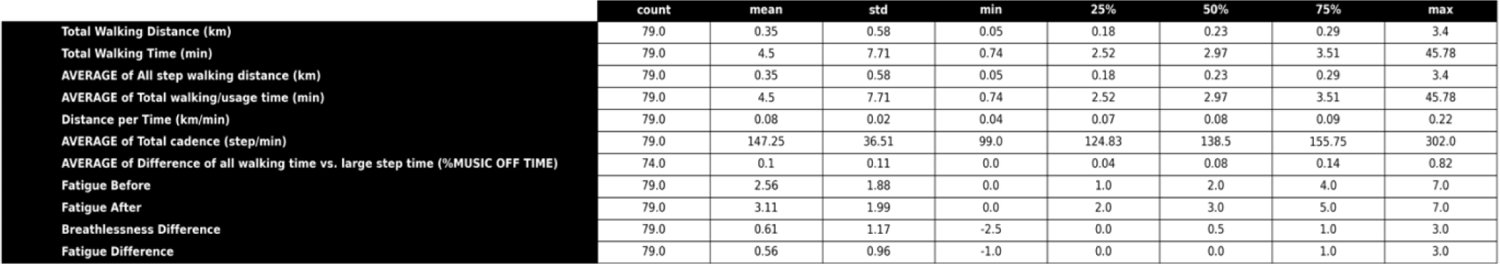
Summary of descriptive statistics for the key variables related to fatigue and breathlessness scores.

|  | count | mean | std | min | 25% | 50% | 75% | max |
| --- | --- | --- | --- | --- | --- | --- | --- | --- |
| Total Walking Distance (km) | 79.0 | 0.35 | 0.58 | 0.05 | 0.18 | 0.23 | 0.29 | 3.4 |
| Total Walking Time (min) | 79.0 | 4.5 | 7.71 | 0.74 | 2.52 | 2.97 | 3.51 | 45.78 |
| AVERAGE of All step walking distance (km) | 79.0 | 0.35 | 0.58 | 0.05 | 0.18 | 0.23 | 0.29 | 3.4 |
| AVERAGE of Total walking/usage time (min) | 79.0 | 4.5 | 7.71 | 0.74 | 2.52 | 2.97 | 3.51 | 45.78 |
| Distance per Time (km/min) | 79.0 | 0.08 | 0.02 | 0.04 | 0.07 | 0.08 | 0.09 | 0.22 |
| AVERAGE of Total cadence (step/min) | 79.0 | 147.25 | 36.51 | 99.0 | 124.83 | 138.5 | 155.75 | 302.0 |
| AVERAGE of Difference of all walking time vs. large step time (%MUSIC OFF TIME) | 74.0 | 0.1 | 0.11 | 0.0 | 0.04 | 0.08 | 0.14 | 0.82 |
| Fatigue Before | 79.0 | 2.56 | 1.88 | 0.0 | 1.0 | 2.0 | 4.0 | 7.0 |
| Fatigue After | 79.0 | 3.11 | 1.99 | 0.0 | 2.0 | 3.0 | 5.0 | 7.0 |
| Breathlessness Difference | 79.0 | 0.61 | 1.17 | -2.5 | 0.0 | 0.5 | 1.0 | 3.0 |
| Fatigue Difference | 79.0 | 0.56 | 0.96 | -1.0 | 0.0 | 0.0 | 1.0 | 3.0 |

**Figure 1.**
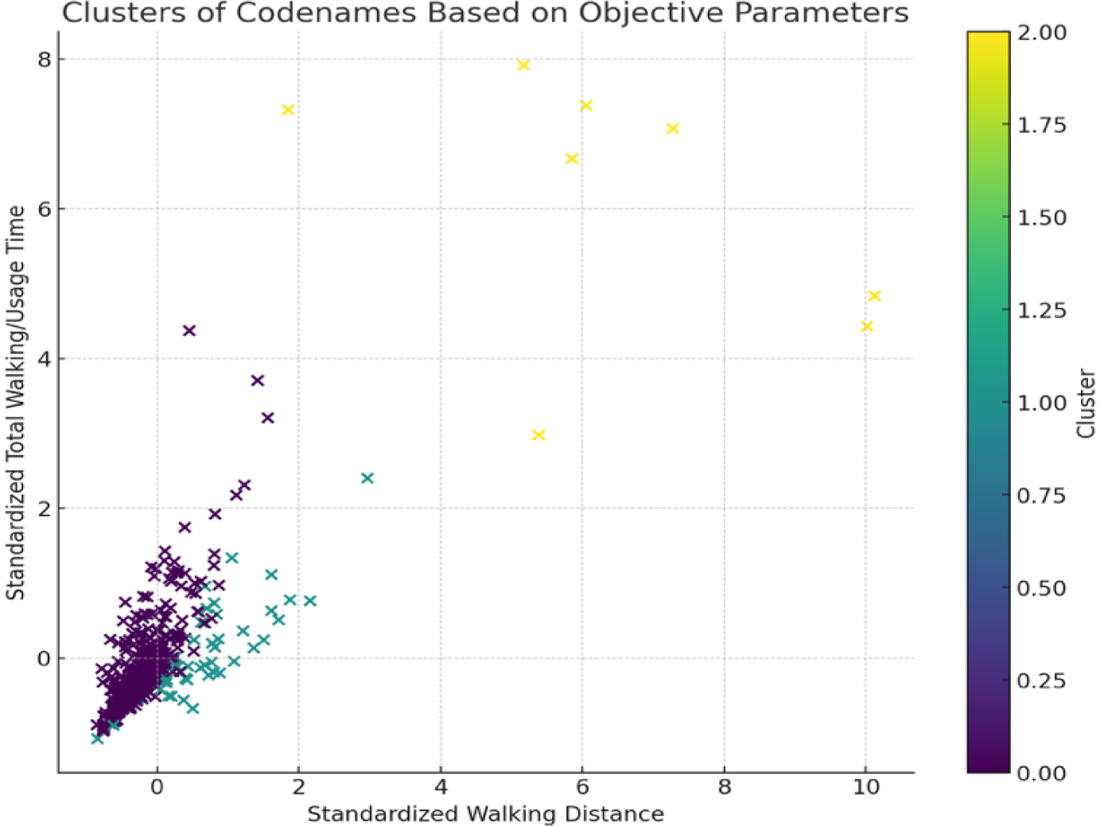
Scatter plot showing the lack of similarities among participant codenames that contributed the data points of objective measurements in the dataset.

### Objective Variable Analysis

#### Data validation

We first used cluster analysis based on user codenames to verify that our dataset is not distorted as a result of the presence of concentrated data from the same participants. The clustering based on the codenames (which also include the stepping sessions or playlist names) revealed very low similarities and no clusters could be identified. For example, for Cluster 0 the user codename numbers are [2, 10, 11, 13, 15], for Cluster 1 they are [0, 1, 3, 8, 12], for Cluster 2 the user codenames are: [4, 5, 6, 7, 9].

### Objective Variable Analysis

Histograms of variable distribution show that most participants walked shorter distances, with a few outliers walking significantly longer distances. Similar to the walking distance, the walking time distribution is also right-skewed, showing that most participants had shorter walking times, with a minority engaging in much longer sessions. Distance per time (km/min) distribution suggests most participants maintained a moderate pace, with fewer participants achieving higher efficiency rates. Average of total cadence (step/min) rates shows a broad range but is less skewed than distance and time, indicating a diverse set of pacing strategies among participants. These visualizations suggest a concentration of participants towards lower values with fewer high-performing outliers.

### Parameter predictions

Measurements of a collection of gait movement parameters are expected to exhibit certain degree of internal heterogeneity even for data samples derived from a single joint and a single sensor output.

**Figure 2.**
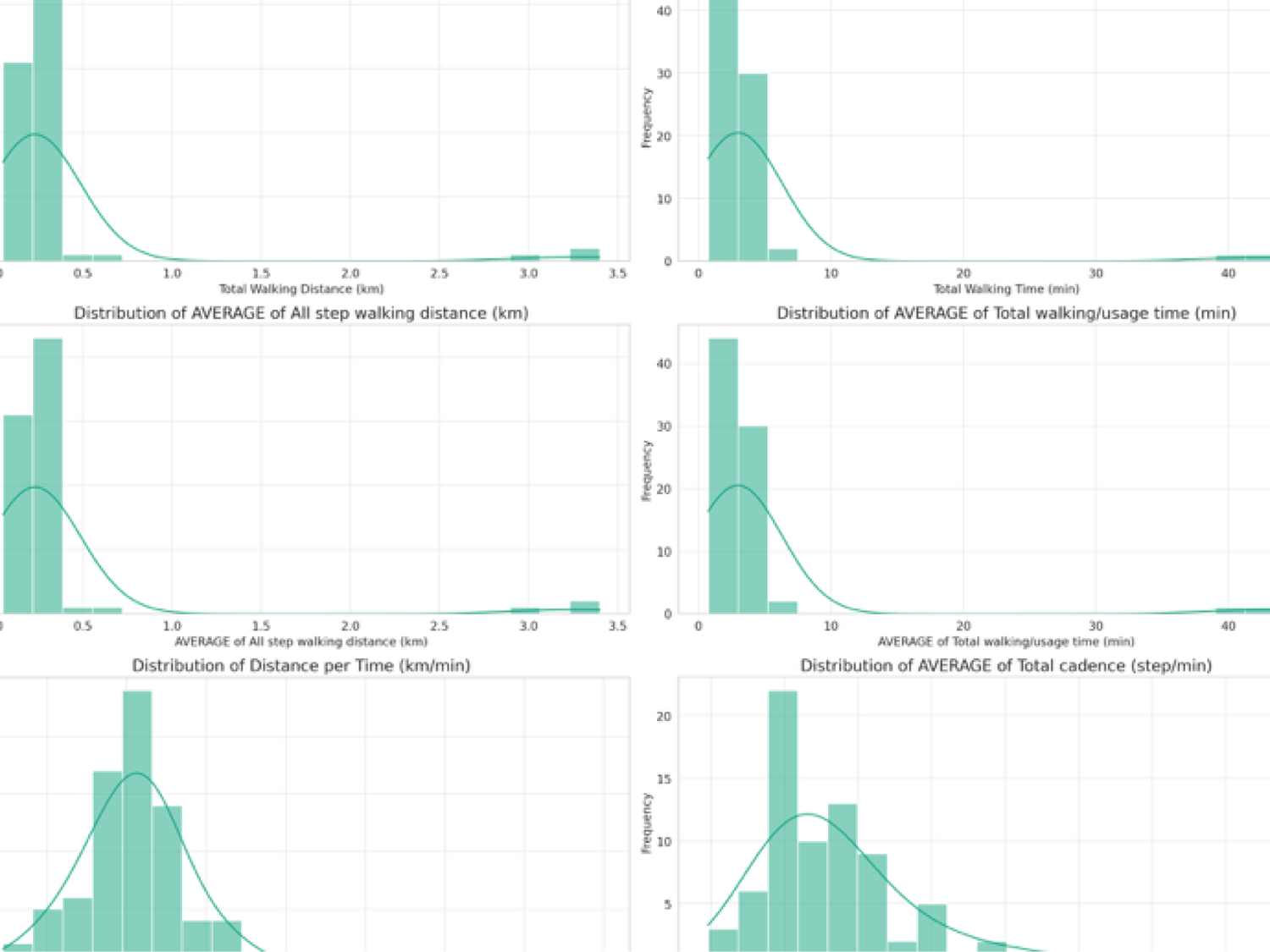
Histograms illustrating the distribution of objective parameters from the SIP exercise data.

**Figure 3.**
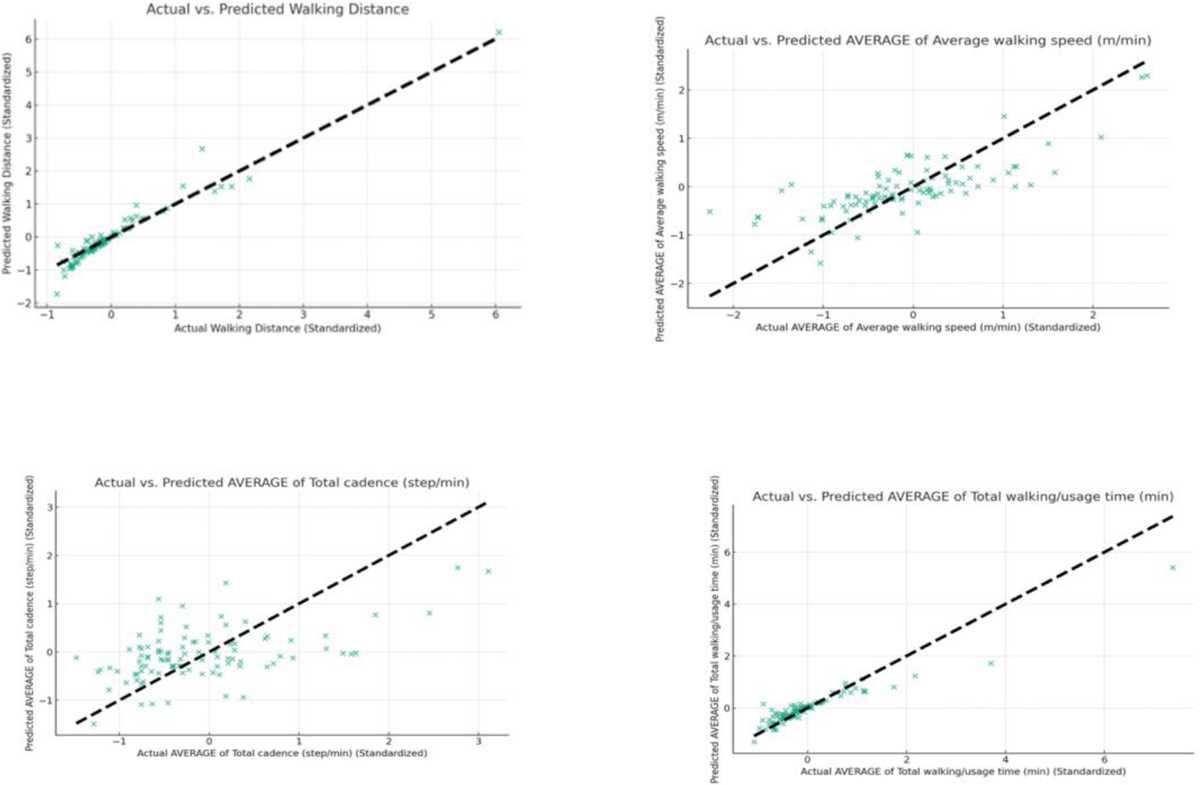
Predicted versus actual plot illustrating the parametric predictability of the regression model described in the text.

To demonstrate this we first asked GPT to generate a predictive regression model of objective walking variables by using the “average of all step walking distance (km)” (standardized) as the dependent variable and other objective parameters as independent variables. We then used the model and the “predicted versus actual plot” to demonstrate the linearity and correlation between the two for each gait parameter. For example, for walking distance, we obtained a R-squared 0.814 indicating that approximately 81.4% of the variance in the walking distance is explained by the regression model.

For the total walking/usage time (min) the obtained MSE = 0.147, indicating a relatively good fit, i.e, the features used in the model are effective in predicting walking/usage time. For average walking speed (m/min), we obtained a MSE = 0.310, showing a moderate fit. This suggests that while the model captures some aspects of walking speed, there may be additional factors not included in the model that influence this parameter. For total cadence (step/min) the MSE = 0.533, indicating the regression model’s fit for predicting cadence is less accurate compared to the other parameters. This suggests that cadence may be influenced by factors not captured by the remaining objective parameters.

### Correlation matrix

The heatmap below illustrates the correlation matrix among the objective parameters. There are strong positive correlations between many of the parameters, as one might expect given their nature. For instance, total walking distance is highly correlated with the average of all step walking distance, which is logical since both metrics measure the distance covered during a particular song. It also suggests that participants were unlikely stepping to duplicated song names during the training session confirming the random nature of song exposure.

Similarly, the total walking time and the average of total walking/usage time also show a strong positive correlation, indicating the consistency in how time is recorded and averaged across sessions. Distance per time shows moderate to strong correlations with other variables, suggesting that as participants walk further or for longer times, their efficiency or pace (distance covered per unit of time) also tends to increase. The average of total cadence has positive correlations with other measures, though not as strong as some of the other relationships. This indicates that while faster stepping (higher cadence) is associated with greater distances and longer times, the relationship varies more among participants compared to other metrics.

**Figure 4.**
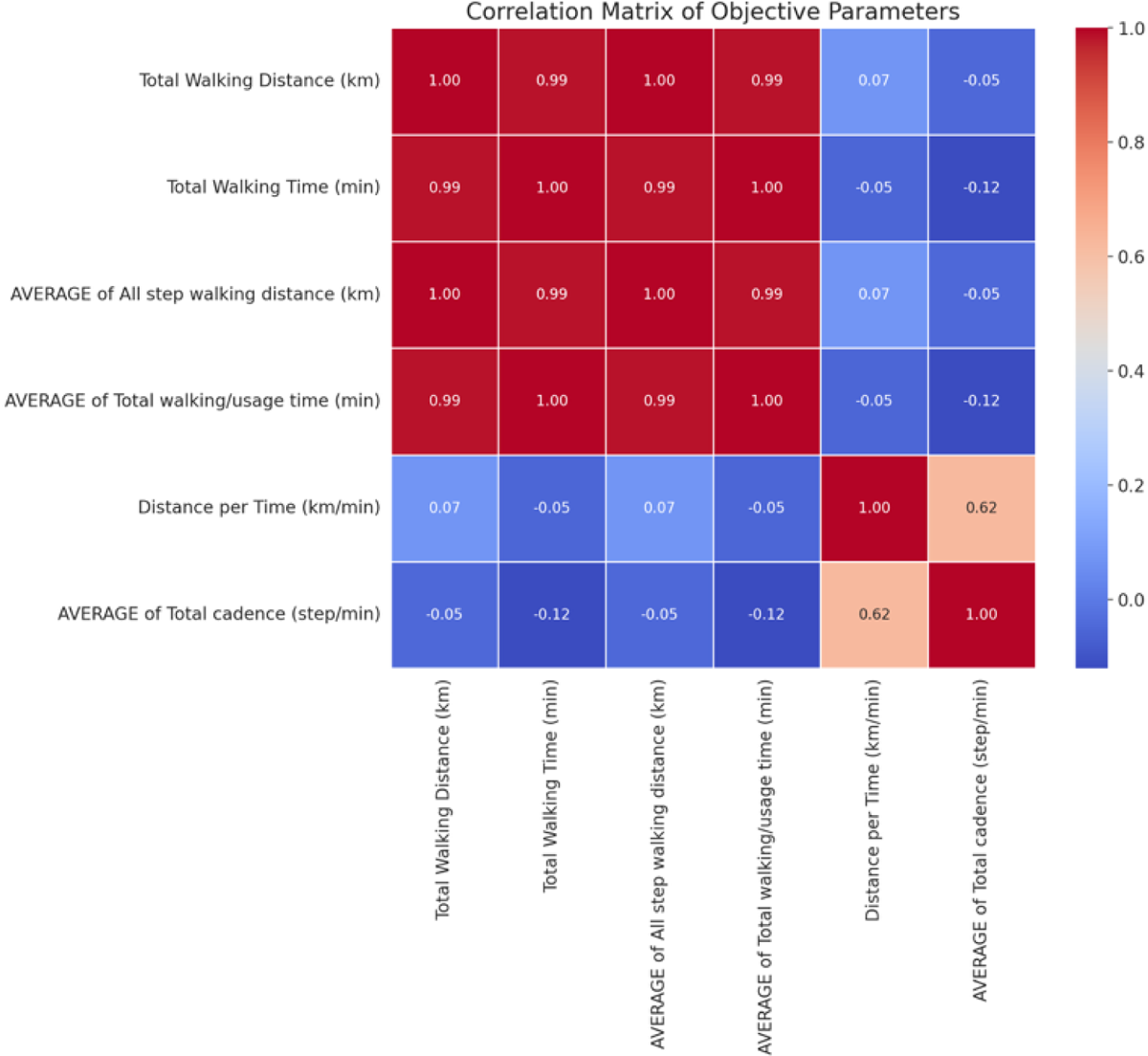
Heatmap illustrating the matrix of correlation coefficients among objective parameters during SIP exercise.

These correlations suggest that the objective measures of physical performance are interrelated, reflecting the complex nature of physical activity during the stepping exercise. Higher performance in one metric often goes hand in hand with higher performance in others, though the strength of these relationships varies.

### Cluster Analysis

The figure below shows the results of the K-means clustering analysis based on the objective parameters: “Average Walking Distance (km)” and “Average Total Walking/Usage Time (min).” The data points are coloured and styled according to their cluster of assignment, with the clusters represented as follows:

**Figure 5.**
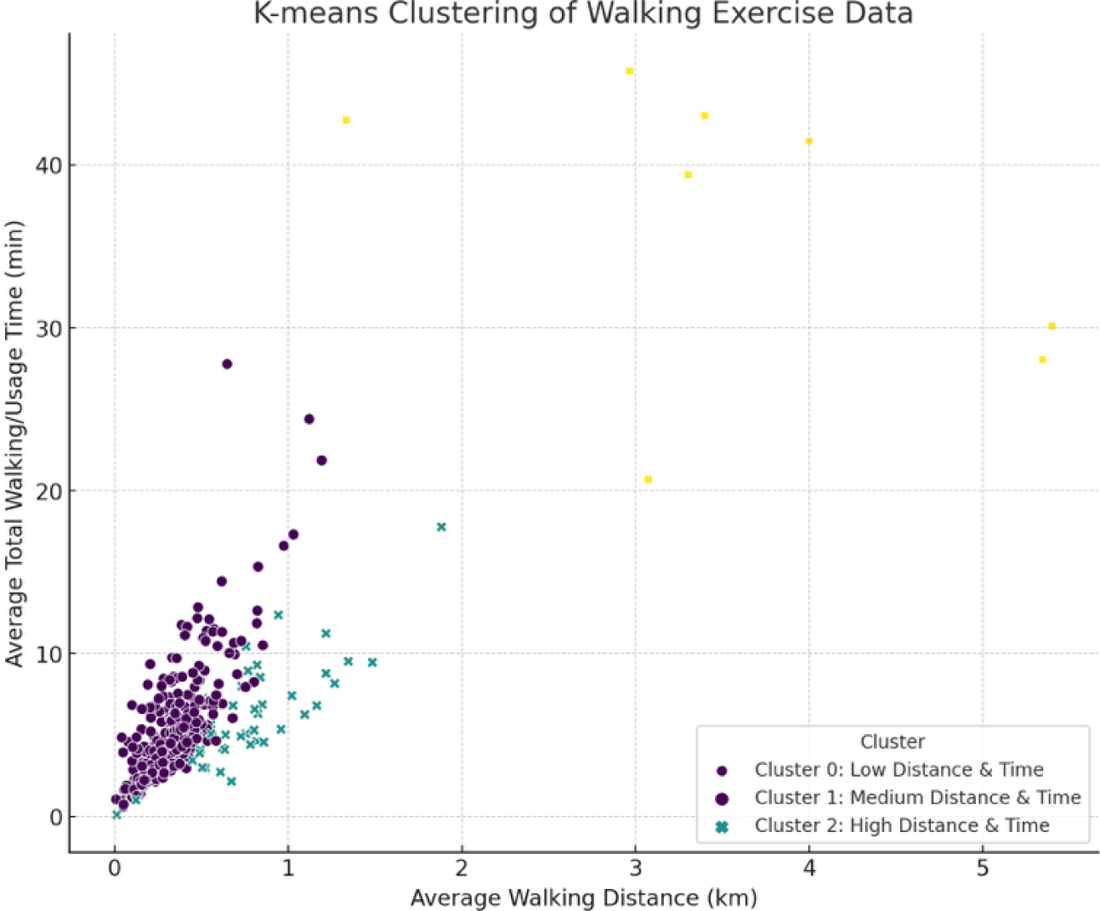
K-means clustering analysis results based on the objective parameters “Average Walking Distance (km)” and “Average Total Walking/Usage Time (min).”

The figure above shows the results of the K-means clustering analysis based on the objective parameters: “Average Walking Distance (km)” and “Average Total Walking/Usage Time (min).” The data points are colored and styled according to their cluster of assignment, with the clusters represented as follows:

– Cluster 0 (Low Distance & Time): This cluster includes participants with lower average walking distances and usage times, suggesting a group with shorter, possibly less intense walking exercises.
– Cluster 1 (Medium Distance & Time): Participants in this cluster have medium ranges of walking distances and usage times, indicating a moderate level of exercise intensity.
– Cluster 2 (High Distance & Time): This cluster represents participants with higher walking distances and longer usage times, suggesting a group engaged in longer or more intense walking exercises.

Two methods were further used for determining the optimal number of clusters for the K-means clustering analysis of the objective parameters:

**Figure 6.**
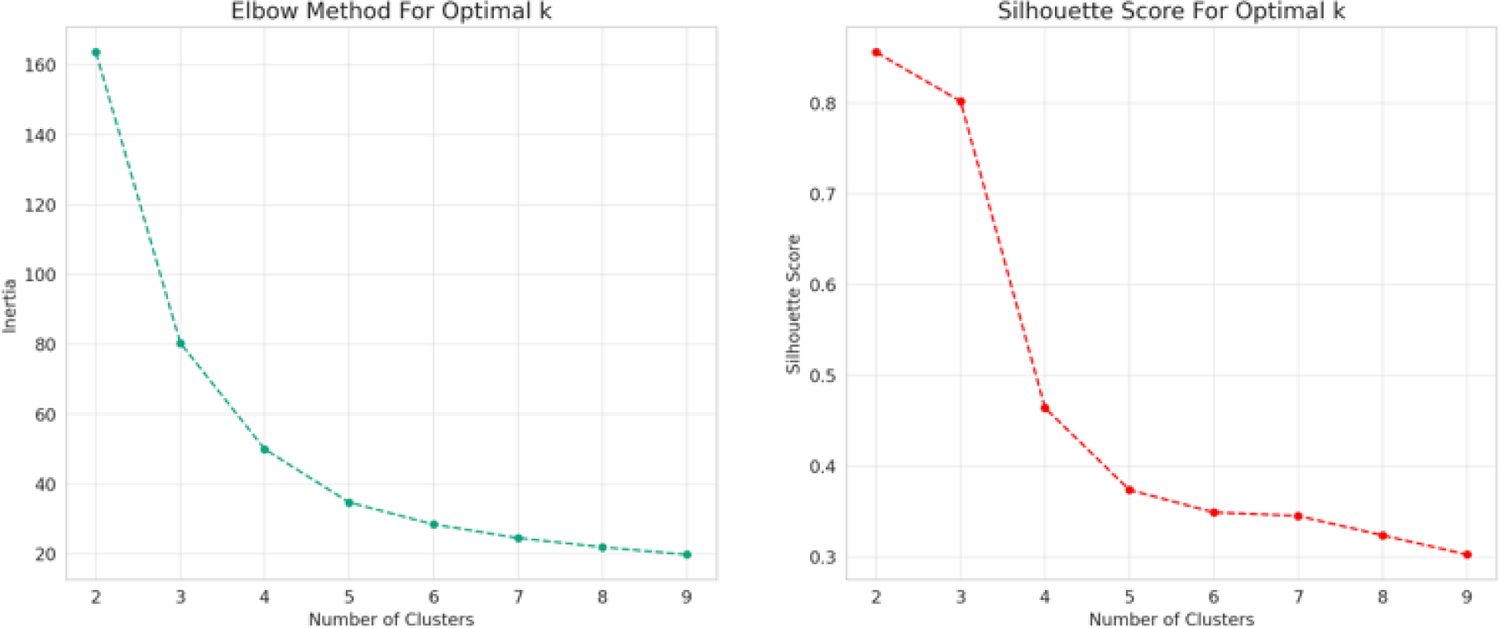
Elbow Method for determining the optimal number of clusters in K-means clustering analysis of objective parameters. Silhouette Score analysis for determining the optimal number of clusters in K-means clustering analysis of objective parameters.

1. Elbow Method: The plot shows the inertia (within-cluster sum of squares) against the number of clusters. The “elbow” point is where the reduction in inertia starts to slow down, indicating an optimal balance between the number of clusters and the compactness of the clusters. From the plot, there’s an elbow around 3 or 4 clusters, suggesting that 3 clusters could be a good choice.
2. Silhouette Score: This score measures how similar an object is to its own cluster compared to other clusters. The plot of silhouette scores against the number of clusters indicates that the score tends to be higher with 2 clusters, with another potential peak at 3 clusters. A higher silhouette score suggests a better-defined cluster structure.

The clustering analysis with 3 clusters revealed distinct groups of participants based on their objective performance metrics during the stepping exercise. Cluster 0 appears to represent a group with moderate physical performance, showing an average walking distance of 0.23 km, walking time of about 3 minutes, and a cadence rate of 144 steps/min. The distance per time for this cluster is 0.08 km/min, suggesting a moderate pace. Cluster 1 seems to consist of participants who engaged in longer and possibly less intensive sessions, with the highest averages in walking distance (3.22 km) and time (42.75 minutes) but a lower average cadence of 126 steps/min. The distance per time is also slightly lower at 0.08 km/min, indicating a steady but less vigorous pace compared to Cluster 2. Cluster 2 is characterized by significantly higher performance in terms of efficiency, with the highest distance per time ratio at 0.20 km/min and an exceptionally high cadence of 294 steps/min. This group, however, has an average walking distance (0.57 km) and time (2.86 minutes) that suggest short, highly intense exercise bursts.

Given the noticed extreme values of cadence by GPT during analysis we asked GPT to re-conduct the clustering analysis by selecting and removing the participants’ data of cadence outliers which is defined as values that lie beyond 1.5 times the interquartile range (IQR) above the third quartile or below the first quartile of cadence data. Even with the refinement of the data by outlier removal, the essence of the cluster distinctions and cluster’s activity profile as described above remains the same::

Cluster 0: Characterized by a Moderate performance profile, this group has longer walking sessions on average, with a moderate pace and cadence rate. Their activities suggest a balanced, sustained effort over time rather than short bursts of high intensity.

Cluster 1: This group exhibits a High performance profile in terms of cadence, indicating more vigorous activity during shorter sessions. Their higher cadence rate, despite the similar distance covered to Cluster 0, suggests a focus on more intense, efficient movement.

Cluster 2: Represents an Average performance profile, with participants engaging in activities of moderate intensity. Their walking sessions are similar in duration to Cluster 1 but at a lower cadence rate, indicating a steadier, less intense form of exercise.

It is important to note that the cadence significantly differentiates the clusters, underlining its importance in characterizing the physical activity profiles.

### Significance Tests

ANOVA test was performed to show the differences in means of the objective parameters across the three clusters.

**Table 2.**
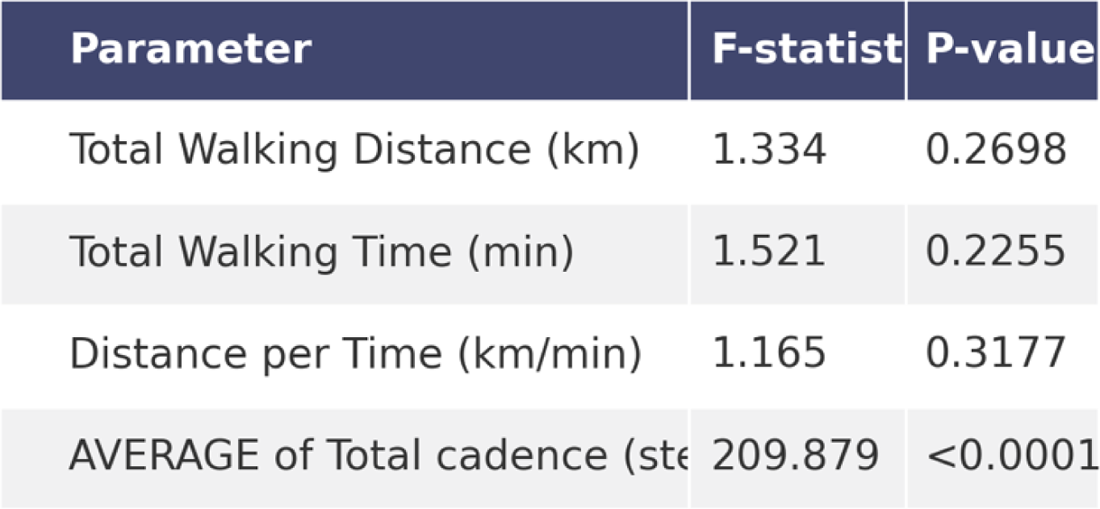
ANOVA analysis showing only cadence is statistically significant different among the means of objective parameters across the three clusters identified through K-means clustering.

This table indicates that the only parameter with a statistically significant difference among the clusters is the “AVERAGE of Total cadence (step/min),” with an extremely low p-value, pointing to significant variance among the groups for this metric. The other parameters—total walking distance, total walking time, and distance per time—did not show statistically significant differences, as indicated by their p-values These results support the distinction made between the clusters, validating the grouping as reflecting differing profiles of physical activity.

#### Xxxx

We also performed additional non-parametric statistical tests to evaluate the differences in objective parameters across the three clusters.

**Table 3.** Results of non-parametric statistical tests assessing the differences in objective parameters across the three clusters identified through K-means clustering.

When dealing with uneven sample sizes across groups, non-parametric tests are often more robust because they don’t assume equal variance or a normal distribution of the data. Kruskal-Wallis test was used for comparing three or more groups while the Mann-Whitney U test was used for comparing two groups at a time. Mood’s Median test was used for assessing whether samples originate from populations with the same median. It’s less sensitive to outliers than mean-based tests and can be a good choice when the data is skewed or has unequal variances. Bootstrapping, although not a test per se, is a powerful resampling method that can estimate the distribution of a statistic (e.g., mean difference between groups) without making assumptions about the population’s distribution. It can be used to construct confidence intervals and perform hypothesis testing, even with uneven group sizes.

The Kruskal-Wallis and Mann-Whitney U tests confirm that the clusters exhibit distinct gait characteristics, with significant differences in walking distances and potentially other related variables.Bootstrapping adds depth by quantifying these differences, particularly highlighting significant disparities or the magnitude of differences in walking distances that could have practical implications (e.g., in therapeutic contexts or fitness assessments). The lack of significant findings from Mood’s median test versus the significant mean differences identified through bootstrapping suggests that while average performances differ markedly, the median (or midpoint) performances among clusters might be less divergent, pointing to skewed distributions or the impact of outliers.

These insights collectively provide a nuanced understanding of how participants grouped into clusters vary in their gait performance, with significant practical differences in walking distances.

### Principal Component Analysis

The PCA analysis revealed that the first three principal components in the dataset can account for over 90% of the total variance in the dataset, suggesting they captured the majority of the information related to physical ability. This can be seen in the plot of cumulative explained variance. This distribution of explained variance indicates that the first three components are sufficient to capture the essential information, reducing the dataset’s complexity while retaining its critical aspects.

**Figure 7.**
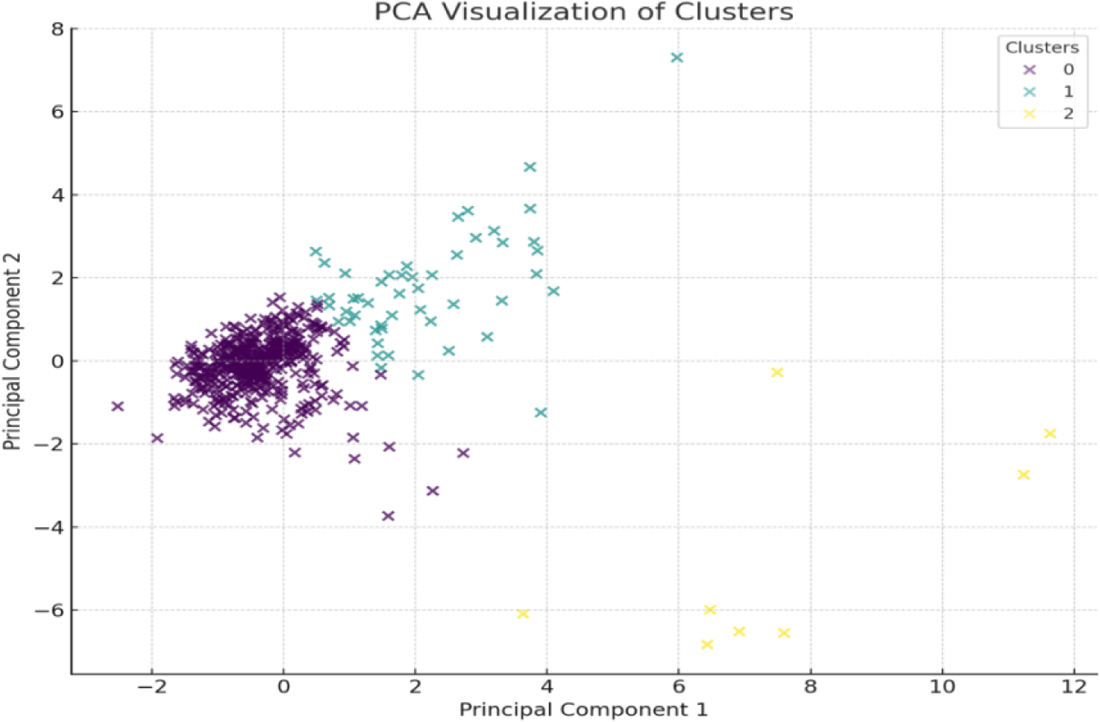
PCA plot indicating the dimensions of the gait parameters can be reduced into two main components.

**Figure 8.**
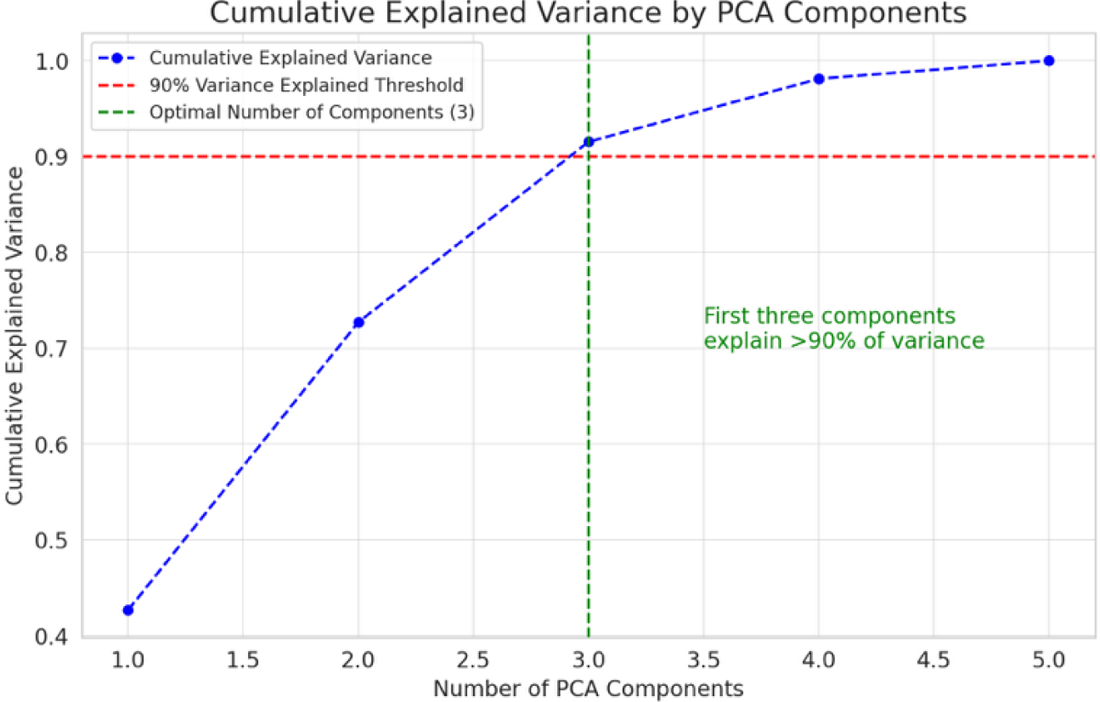
Cumulative explained variance plot indicating the variance of gait parameters can be determined as a function of number of PCA components.

The cumulative explained variance plot further shows that the variance in the dataset exhibits a significant drop-off after the third component with the first component explains approximately 42.65% of the variance, second component for about 30.05% of the variance, third component contributing 18.84% to the variance. The results indicate that a multi-dimensional view of physical ability, encompassing various aspects of gait and movement, can be effectively captured through PCA on the selected objective measurements.

The PCA component loadings were further used to illustrate the contribution of each objective measurement to the first three principal components. The PCA scatter plot visualizes participants in a reduced-dimensional space based on their objective performance metrics, with colors representing the clusters identified earlier showing distinct groupings corresponding to the clusters, indicating that the PCA transformation retains the separation based on physical performance metrics. Principal Component 1 (PC1) and Principal Component 2 (PC2) together capture the variance in the dataset that most significantly contributes to the differences among participants, aligning with the cluster assignments.

**Figure 10.**
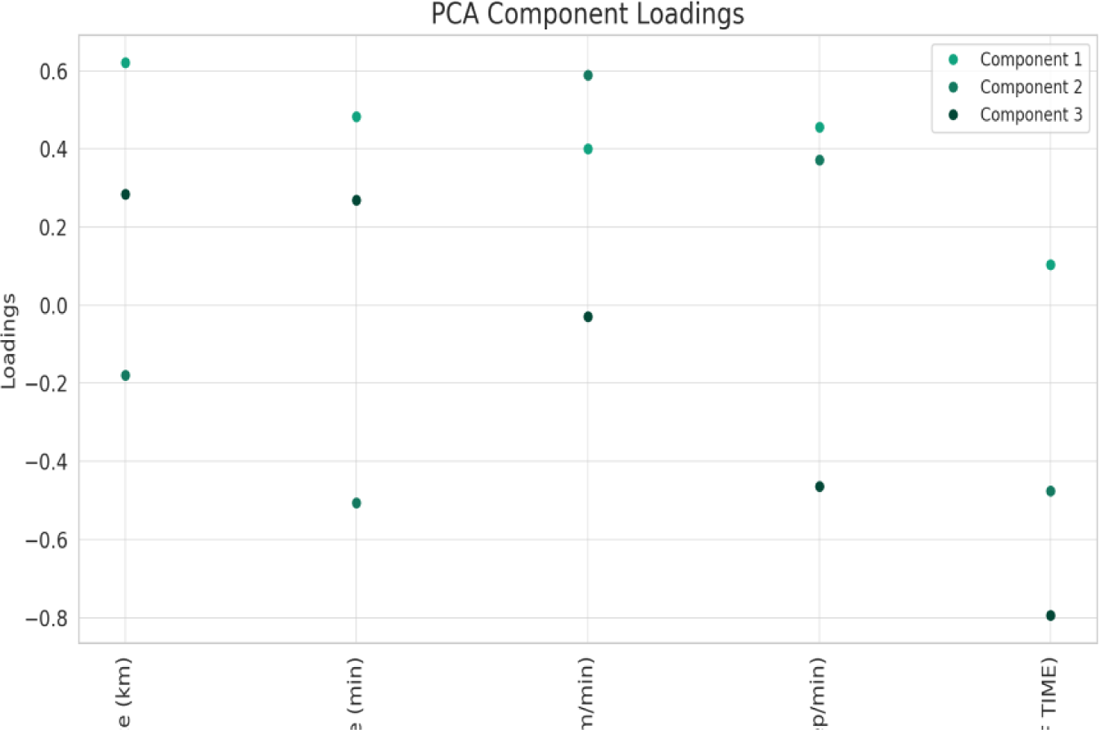
PCA Component Loadings Plot. This plot illustrates the contribution of each objective measurement to the first three principal components obtained from Principal Component Analysis (PCA).

The Kruskal-Wallis test was performed on the first two principal components (PC1 and PC2) scores across the clusters as shown in the table below.

**Table 5.**
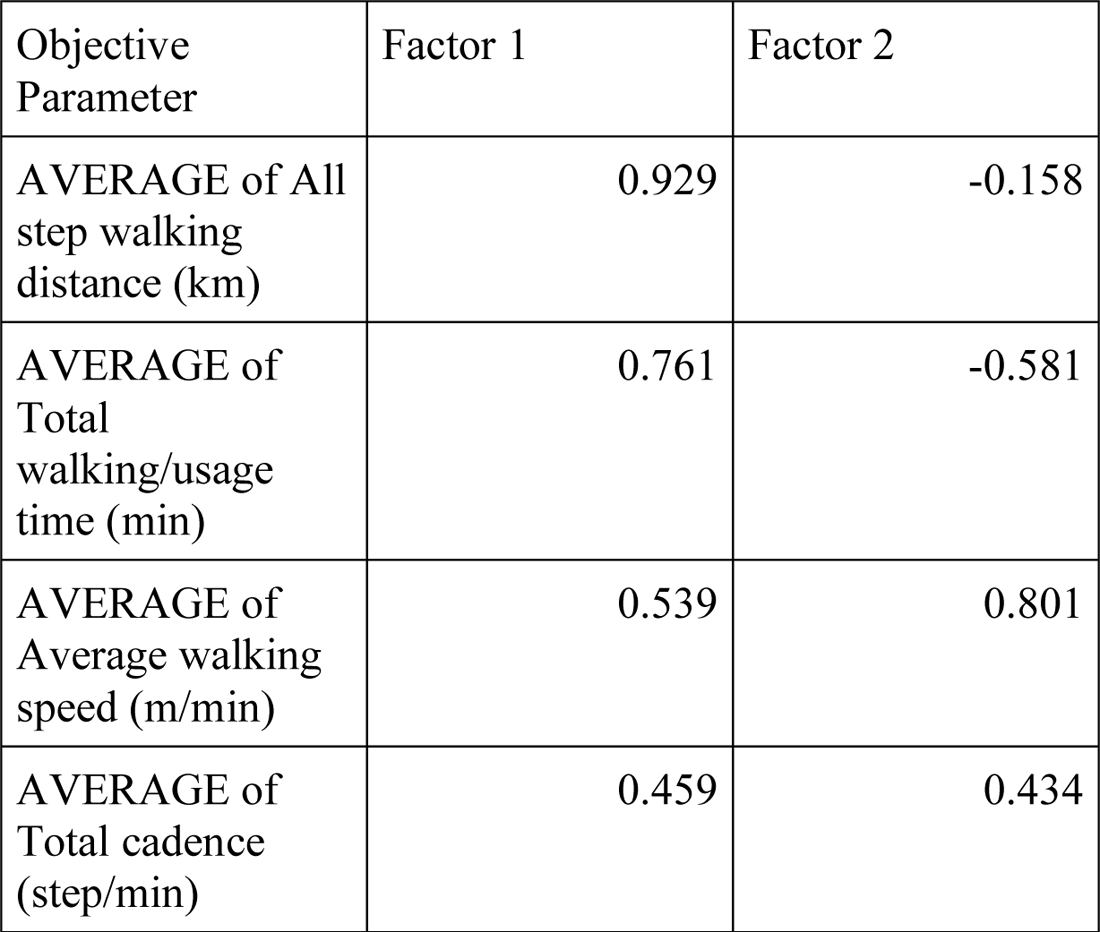
Kruskal-Wallis Test Results for PC1 and PC2 Scores Across Clusters.

| Objective Parameter | Factor 1 | Factor 2 |
| --- | --- | --- |
| AVERAGE of All step walking distance (km) | 0.929 | -0.158 |
| AVERAGE of Total walking/usage time (min) | 0.761 | -0.581 |
| AVERAGE of Average walking speed (m/min) | 0.539 | 0.801 |
| AVERAGE of Total cadence (step/min) | 0.459 | 0.434 |

We found that for PC1 (p=0.826), there is no statistically significant difference in the distribution of PC1 scores across the clusters. This indicates that, based on PC1, the physical abilities in terms of the variances of objective parameters among participants *within* different clusters are relatively similar. Likewise, for PC2 (P=0.116) there is no statistically significant difference in the distribution of PC2 scores across the clusters. This suggests that the participants’ physical abilities, as captured by the first two principal components of variances, are not significantly different across the clusters identified in the PCA analysis, despite the fact that the mean or median values of the same variable set are dissimilar as revealed by K-mean clustering (see above).

### Hierarchical Clustering Analysis

This analysis was used to understand the natural groupings in the dataset further and how closely related the participants are within and across the clusters identified by K-means. We’ll create a dendrogram to illustrate this. We used distance metrics derived from the PCA-transformed features to further confirm the optimal number of clustering described above.

In the figure, each leaf (or vertical line) represents a participant. The height of the horizontal lines joining two leaves or clusters represents the distance or dissimilarity between them. The lower the height, the more similar the participants or groups. Color changes in the dendrogram indicate different clusters based on the specified color threshold, which helps visualize how participants group together naturally before reaching the threshold. The dendrogram supports the cluster analysis by showing the natural groupings within the dataset and how individuals or clusters merge at various levels of similarity.

**Figure 11.**
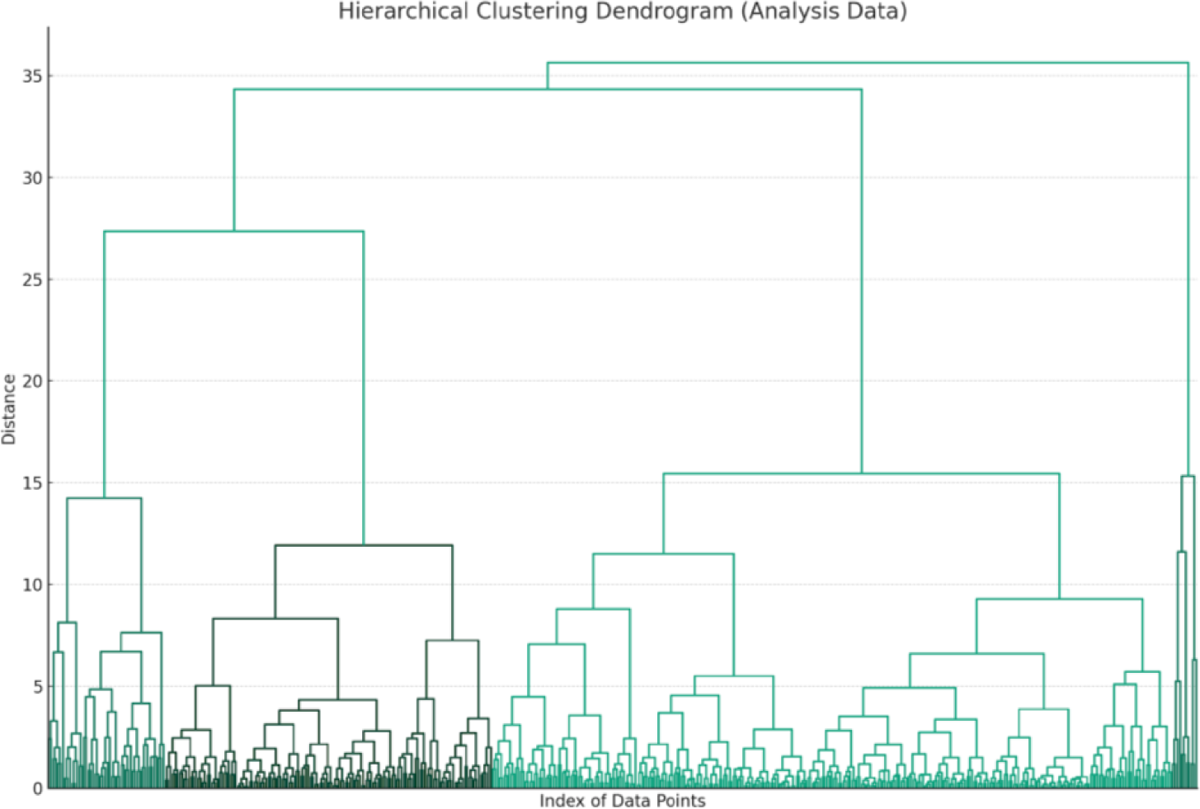
Dendrogram illustrating the results of the Hierarchical Clustering Analysis. In this visualization, each leaf represents a participant.

### Factor Analysis for Validation

Factor analysis was applied to further explore and validate the underlying dimensions of the objective parameters, which can provide additional insights into the data structure and support the conclusions drawn from clustering. The factor analysis on the objective parameters is implemented as factor loadings, which represent the correlations between each variable and the factors. These loadings help identify the underlying dimensions (factors) that explain the observed correlations and relative importance of among the variables used for clustering analysis.

Factors 1 and 2 are strongly correlated with the total walking distance, total walking time, average step walking distance, and average total walking/usage time. This suggests that these two factors capture aspects related to the overall extent and duration of the exercise. Factor 1 appears to equally represent distance and time, indicating it might capture the overall activity level. Factor 2 shows a differentiation between distance/time measures and efficiency (distance per time), suggesting it might capture aspects related to exercise efficiency or intensity. Factor 3 has significant loadings for distance per time and average total cadence but in opposite directions. This factor could represent the trade-off between exercise efficiency and pacing strategy, with higher cadence possibly compensating for lower distance per time efficiency or vice versa.

These findings from the factor analysis provide insights into the underlying structure of the data overall activity Level (duration and extent), efficiency/intensity (differentiated by how efficiently participants perform the exercise and pacing strategy (the balance between maintaining a higher pace (cadence) and the overall efficiency of the exercise.

Factor analysis has thus helped to validate and further understand the distinctions between the clusters identified earlier, emphasizing different aspects of physical performance during the stepping exercise.

**Figure 12.**
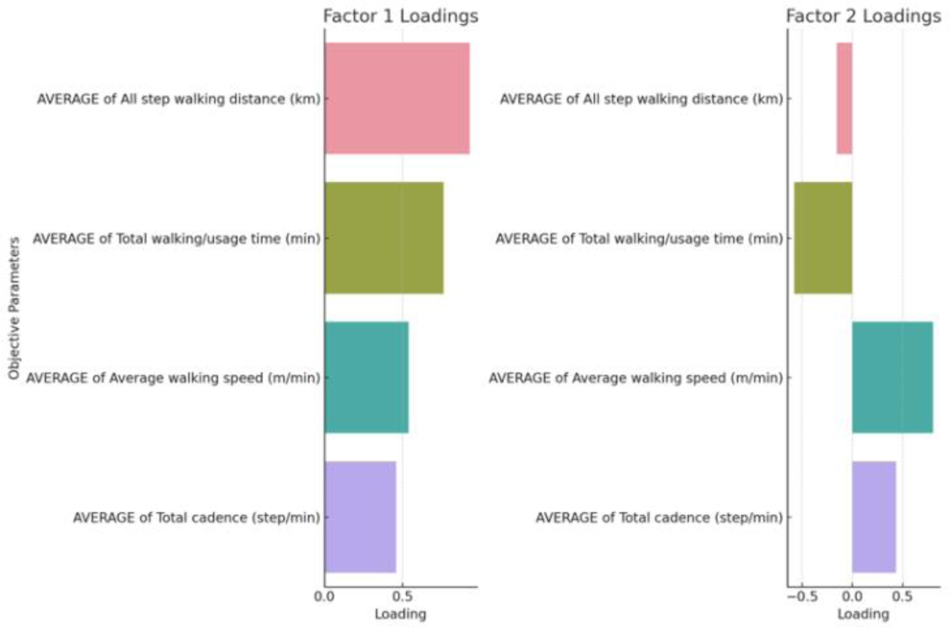
Factor loadings resulting from Factor Analysis on the objective parameters.

**Figure 13.**
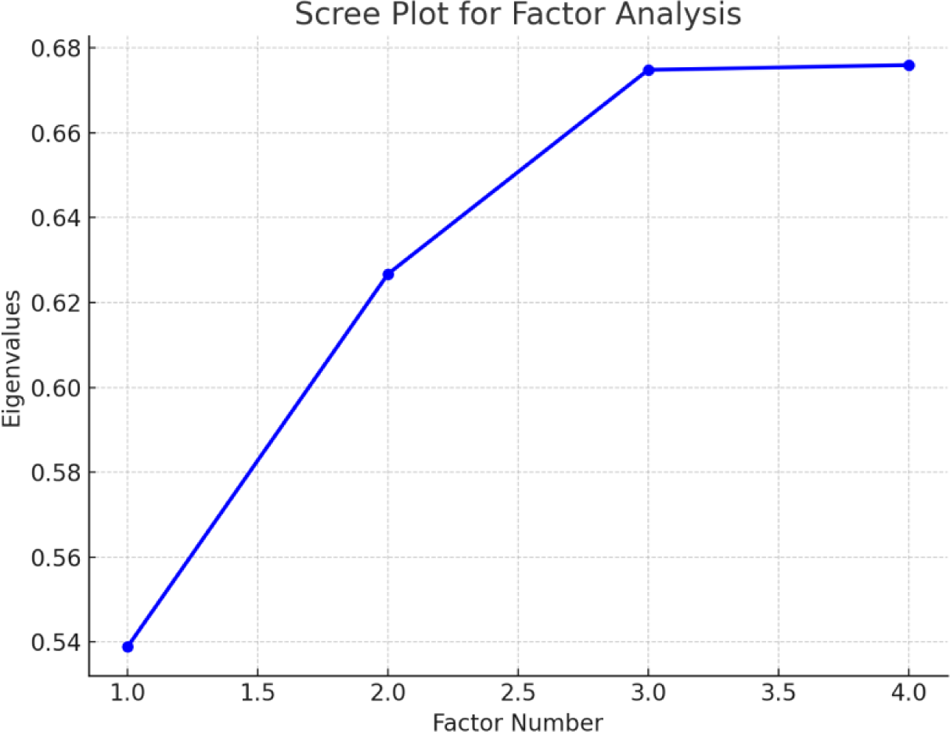
Scree plot identifying latent variables

Finally, we used the scree plot to identify latent variables that explain patterns of correlations among observed variables by determining the number of factors to retain. The results confirm that the three factors mentioned above captured the major values of factor differentiation (Eigenvalues) evidenced by the steep drop in Eigenvalues after the three factor loadings.

### Subjective Variables Analysis

The table below summarizes the descriptive statistics analyzed in the study related to fatigue and breathlessness scores.

|  | count | mean | Std | min | 25% | 50% | 75% | max | mode |
| --- | --- | --- | --- | --- | --- | --- | --- | --- | --- |
| **Fatigue Before** | 79 | 2.56 | 1.88 | 0.00 | 1.00 | 2.00 | 4.00 | 7.00 | 0.00 |
| **Fatigue After** | 79 | 3.11 | 1.99 | 0.00 | 2.00 | 3.00 | 5.00 | 7.00 | 2.00 |
| **Breathlessness**<br>**Difference** | 78 | 0.61 | 1.18 | -2.50 | 0.00 | 0.50 | 1.00 | 3.00 | 0.00 |
| **Fatigue**<br>**Difference** | 79 | 0.56 | 0.96 | -1.00 | 0.00 | 0.00 | 1.00 | 3.00 | 0.00 |

### Relationship of Subjective Score Changes

The graphs below indicate that, comparing to objective parameters, breathless and fatigue score changes show a clear and linear correlation indicating the participants correctly used the both scores for rating exertion.

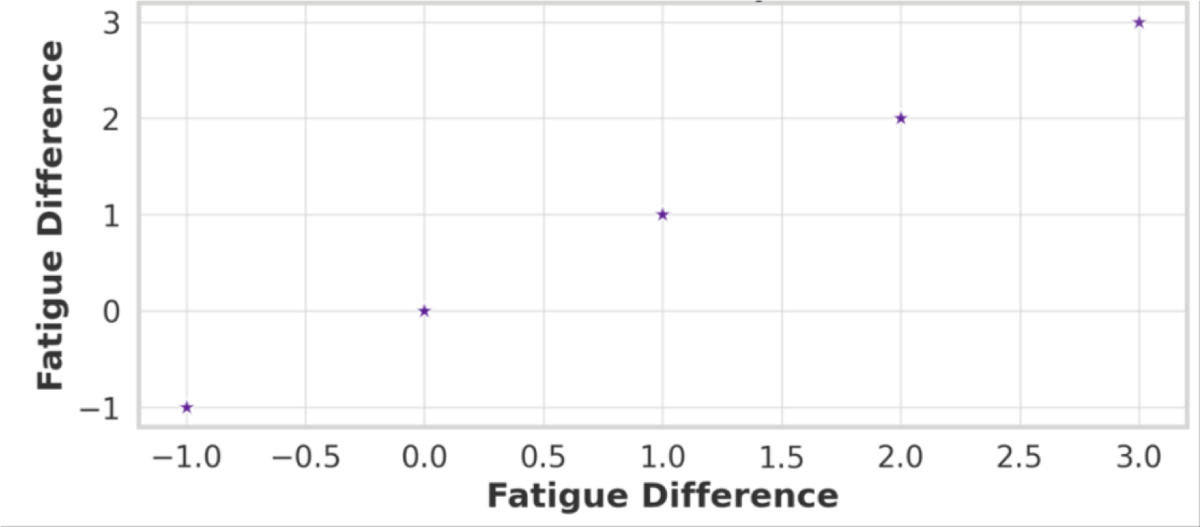

### Clustering Analysis

K-means clustering was applied to categorize participants into three distinct groups based on their differences in subjective fatigue and breathlessness scores. The clusters revealed one group with a moderate increase in scoring, one with minimal changes, and another with significant increases in both fatigue and breathlessness.

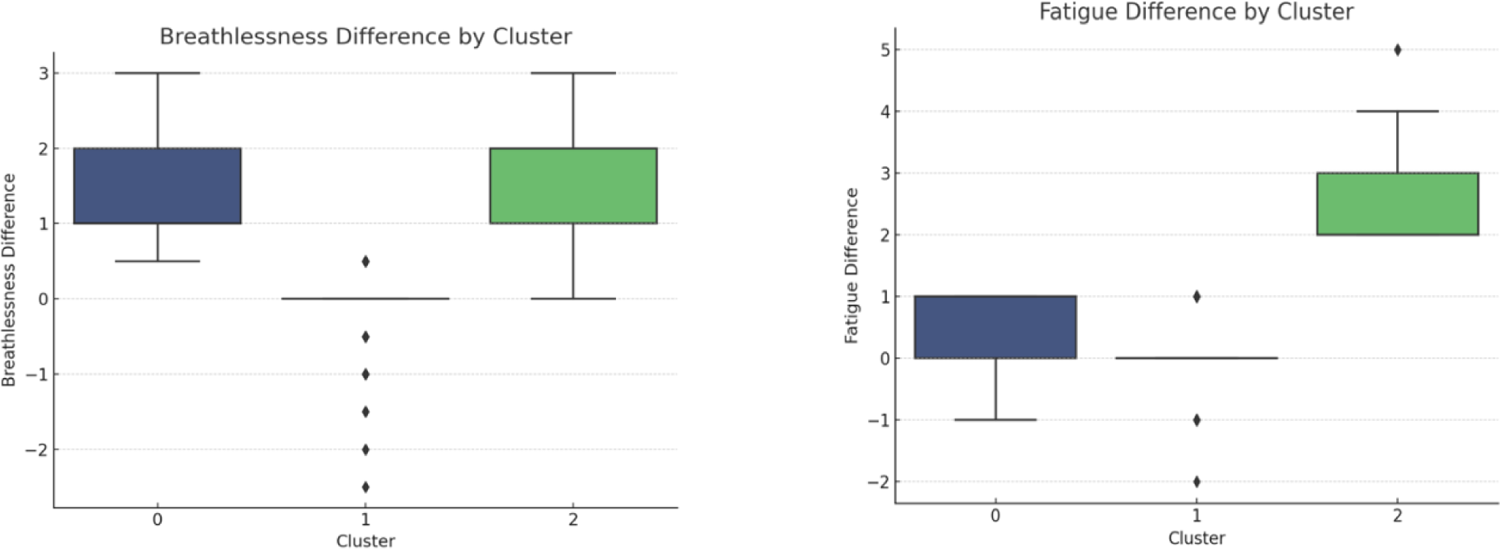

The data highlights the variability in subjective responses to physical activity. This variability underscores the importance of considering individual differences in physical activity recommendations and interventions.

The key statistic features of K-mean clusters are shown below:

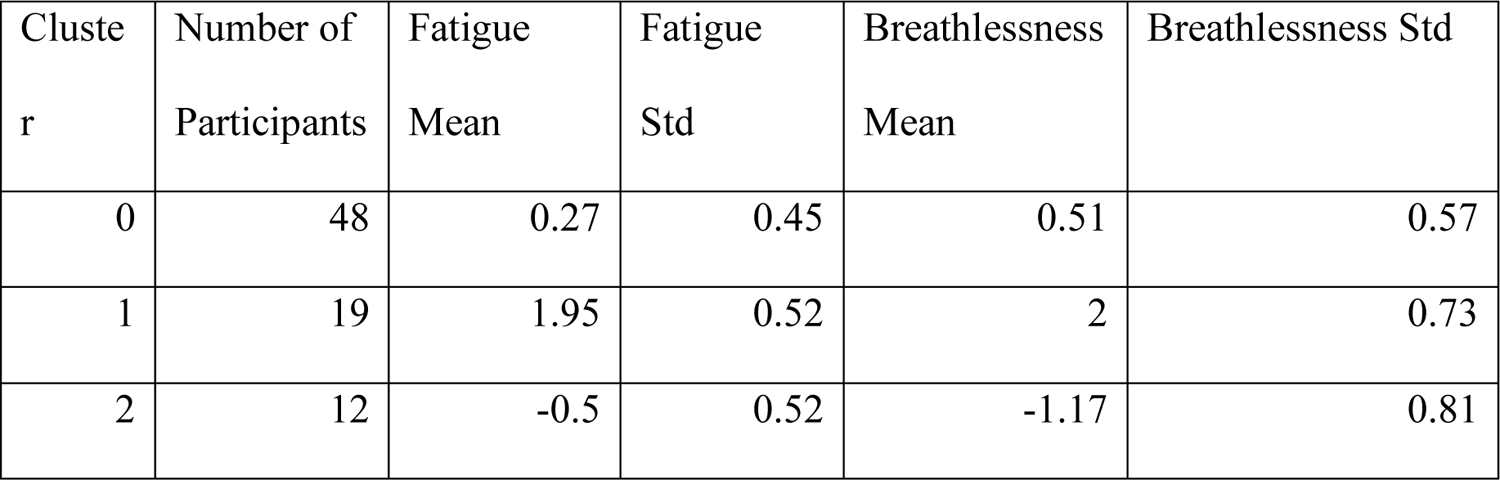

| Cluster | Number of Participants | Fatigue Mean | Fatigue Std | Breathlessness Mean | Breathlessness Std |
| --- | --- | --- | --- | --- | --- |
| 0 | 48 | 0.27 | 0.45 | 0.51 | 0.57 |
| 1 | 19 | 1.95 | 0.52 | 2 | 0.73 |
| 2 | 12 | -0.5 | 0.52 | -1.17 | 0.81 |

Cluster 0 represents the largest group with 48 participants, showing moderate increases in fatigue and breathlessness. Cluster 1 includes 19 participants who, on average, reported the highest increase in both fatigue and breathlessness, suggesting this group found the exercise to be more strenuous. Cluster 2 is composed of 12 participants who, intriguingly, experienced a decrease in both fatigue and breathlessness on average, indicating possible improvements in their physical condition or adaptation to the exercise.

### Statistical Tests

ANOVA test was conducted to examine the statistical significance of the differences among the clusters. The results for both fatigue and breathlessness differences were highly significant (p < 0.05), indicating statistically significant differences among the group means.

| Test | p-value |
| --- | --- |
| ANOVA - Breathlessness Difference | 1.58E-20 |
| ANOVA - Fatigue Change | 3.46892E-24 |
| Kruskal-Wallis - Breathlessness Difference | 7.86526E-12 |
| Kruskal-Wallis - Fatigue Change | 9.8625E-13 |

The statistical significance suggests that the clustering has meaningfully segmented the participants according to their subjective responses to the exercise, supporting the validity of these clusters as distinct groups with differing subjective experiences.

### Scatter Plot of Clustering Analysis

The scatter plot below further illustrates the relationship between the differences in fatigue and breathlessness before and after exercise, categorized by different physical ability levels.

**Figure 17:**
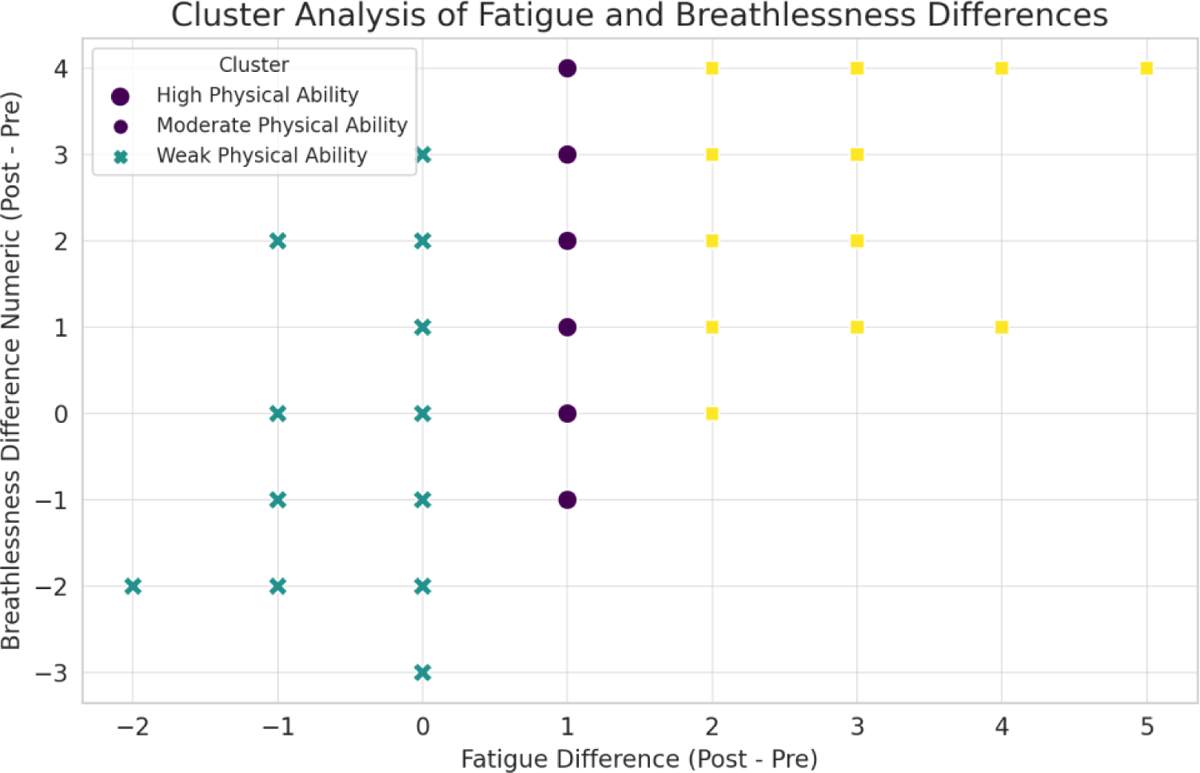
Scatter plot showing the clustering of physical ability based on differences in fatigue and breathlessness.

### Principal Component Analysis

PCA visualization of subjective changes, along with the cluster labels, provides a graphical representation of how the data points (representing participants) are distributed across the two principal components, which together account for a significant portion of the variance in the dataset. Principal Component 1 explains approximately 85.13% of the variance. Principal Component 2 accounts for about 14.87% of the variance. This indicates that most of the variance in the subjective changes can be captured by the first principal component, suggesting that it might represent a dominant pattern in how participants’ subjective experiences of breathlessness and fatigue change due to the exercise. The clusters appear to be well-separated in this reduced-dimensional space, reinforcing the validity of the clustering based on subjective changes. This feature differs from the PCA analysis results on objective parameters where a homogeneity of variance (but not mean or median values) was observed, suggesting PCA can reveal different aspects of objective vs subjective variables when variances are used in grouping participants’ physical abilities.

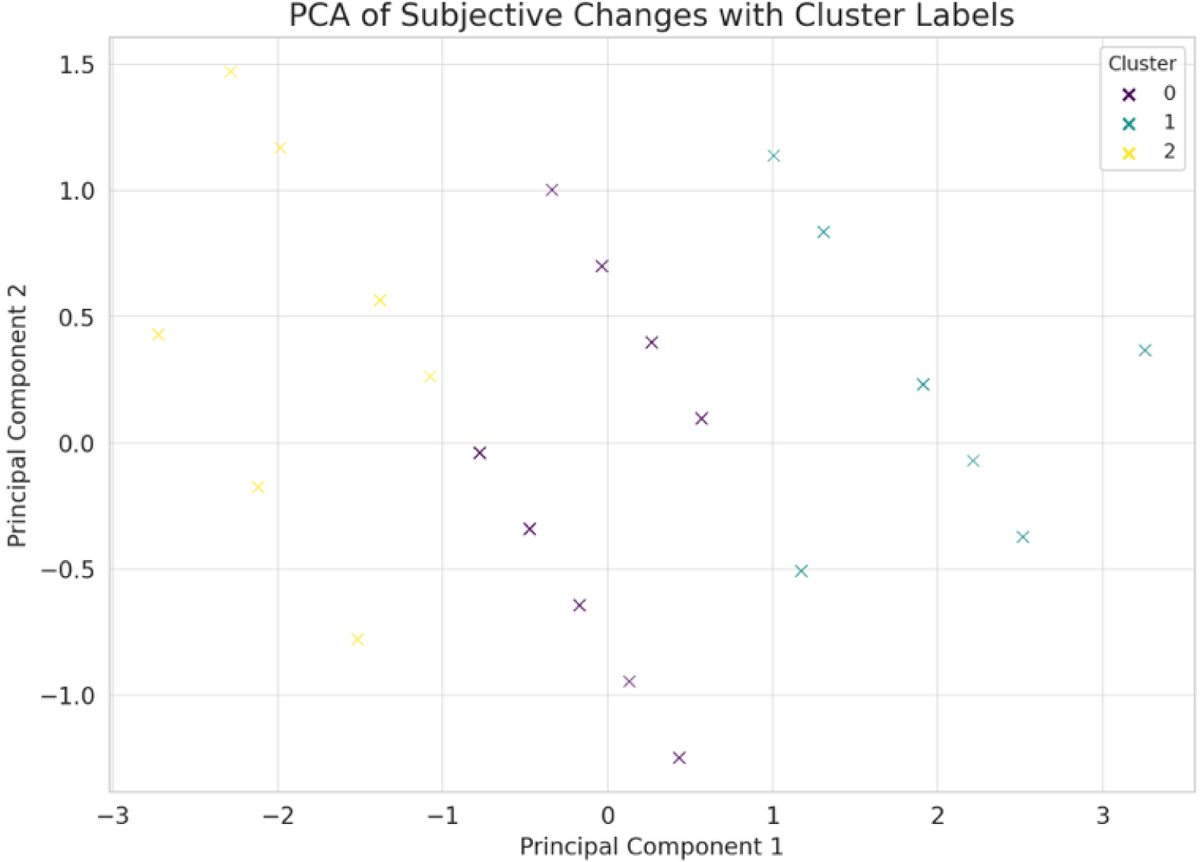

Next, we perform FA to further identify underlying factors that might explain the observed variations in subjective changes. The descriptive statistics table below for the principal components (PC1, PC2, PC3) across subjective physical ability classification groups (High, Moderate, Weak) outlines the mean and standard deviation for each principal component within these groups, offering insights into the central tendencies and variability of the objective measurements captured by PCA.

| Group | Component | Mean | Standard Deviation |
| --- | --- | --- | --- |
| High | PC1 | 0.15 | 1.85 |
| **High** | PC2 | 0.0<br>0 | 1.43 |
| **High** | PC3 | -<br>0.0<br>3 | 1.03 |
| **Moderate** | PC1 | -<br>0.1<br>3 | 1.27 |
| **Moderate** | PC2 | 0.0<br>0 | 1.18 |
| **Moderate** | PC3 | 0.0<br>7 | 0.97 |
| **Weak** | PC1 | -<br>0.0<br>2 | 1.18 |
| **Weak** | PC2 | 0.0<br>0 | 1.05 |
| **Weak** | PC3 | -<br>0.0<br>4 | 0.92 |

### High clustering analysis

The dendrogram from the hierarchical cluster analysis provides a visual representation of the clustering process for the subjective changes in breathlessness and fatigue. In this figure each vertical line represents a participant or a cluster of participants. The height of the horizontal lines joining two clusters or participants indicates the distance or dissimilarity between them. Lower heights indicate more similar participants or clusters. The color threshold indicates the point at which clusters are distinguished. Below this threshold, clusters merge into larger groups, suggesting a natural grouping based on the distance metric used.

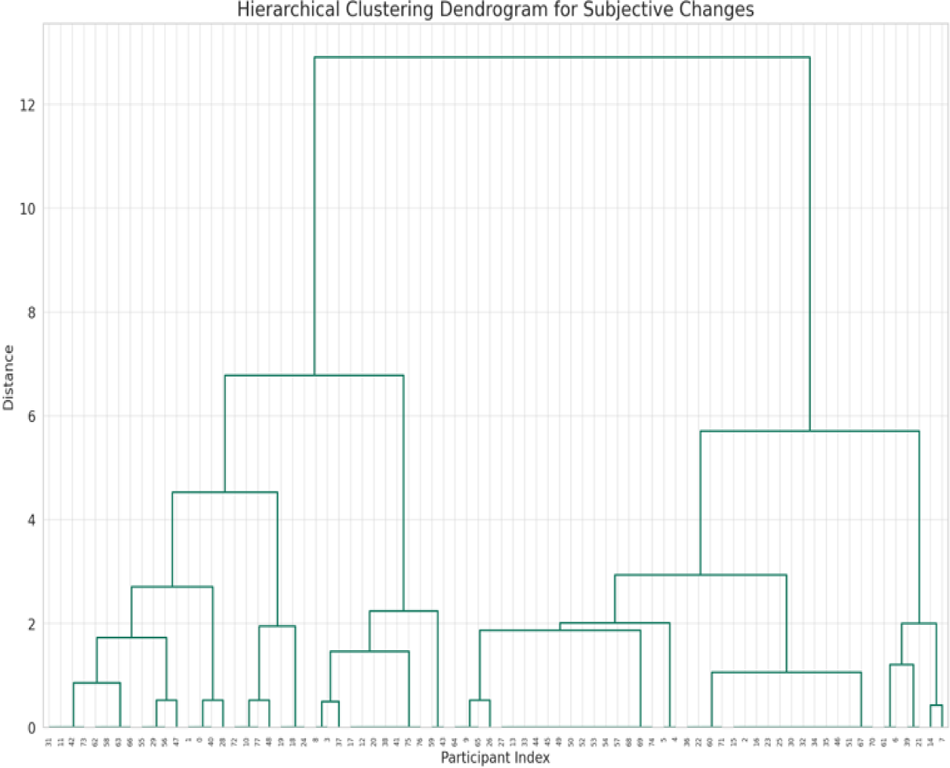

This hierarchical clustering approach offers a nuanced view of how participants group based on their subjective changes, complementing the K-Means clustering results. It allows us to observe the structure and similarity within the data without predefining the number of clusters, providing another layer of validation for the cluster distinctions identified earlier.

### Correlation between subjective classifications of physical ability and objective measurements among all parameters

#### Scatterplots

The figures above show there is no obvious trend of linear relationship between the two categories of exercise variables. The correlation heatmap below further displays the relationships between all numerical parameters in the dataset. Each cell in the heatmap provides the correlation coefficient between the variables on the x and y axes, ranging from - 1 (perfect negative correlation) to +1 (perfect positive correlation), with 0 indicating no correlation. This visualization helps in identifying which variables have strong associations, either positive or negative, with each other.

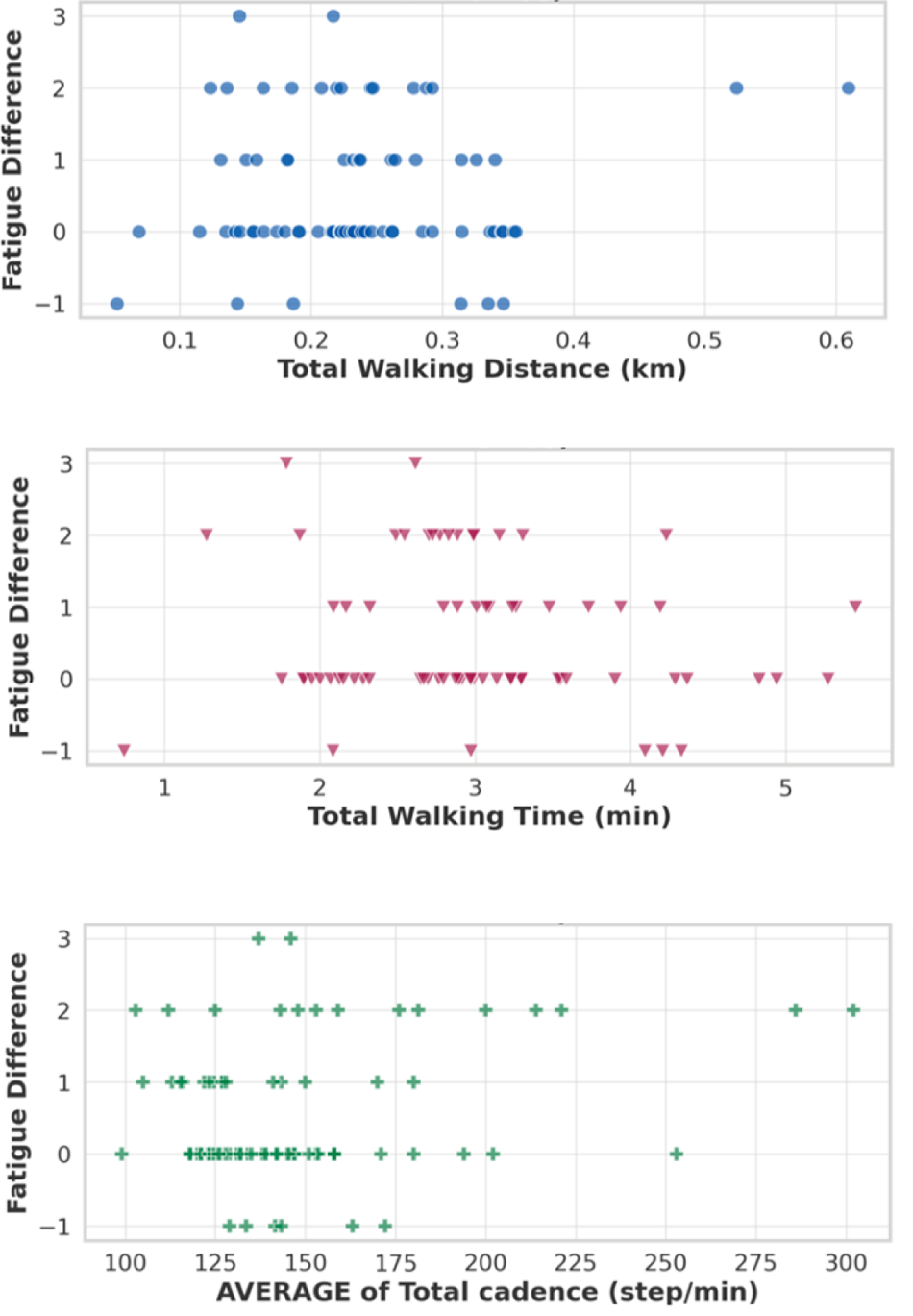

The top three strongest correlations, excluding self-correlations, are:

1. Between ‘Fatigue Before’ and ‘Fatigue After’ with a correlation coefficient of approximately 0.845.
2. Between ‘Average of Total walking/usage time (min)’ and ‘Average of All step walking distance (km)’ with a correlation coefficient of approximately 0.799.
3. Between ‘Breathlessness After Numerical’ and ‘Fatigue After’ with a correlation coefficient of approximately 0.631.

These pairs show the highest degree of association in the dataset according to the correlation analysis. In contrast, we found there are very weak relationships between the total walking/usage time and the fatigue scores both before and after the activity, as well as the difference in fatigue scores. Similarly, weak correlations were observed with breathlessness scores, suggesting that other factors not captured by these parameters may play a significant role in determining subjective experiences of fatigue and breathlessness.

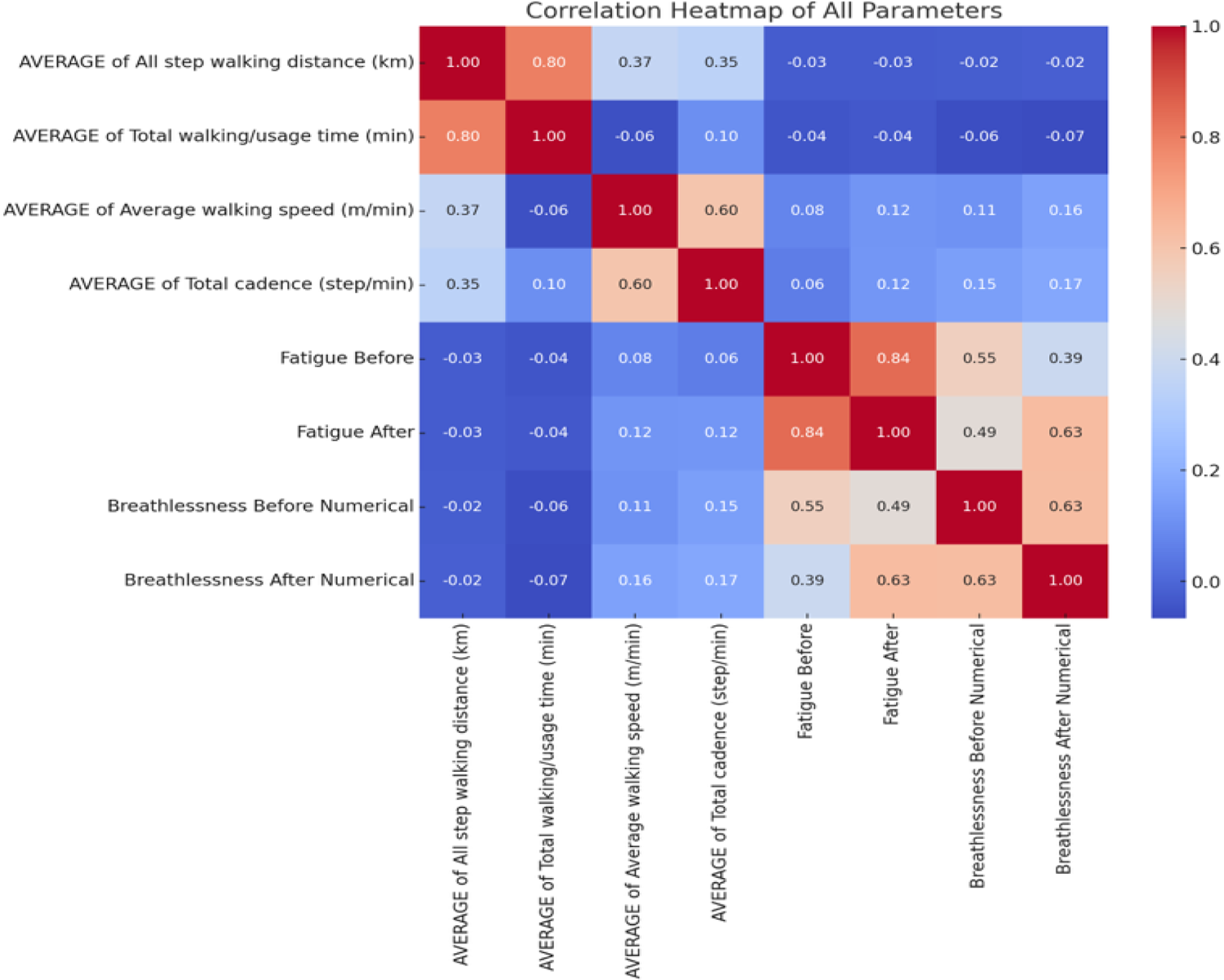

The Spearman correlation analysis between “Total Walking Distance (km)” and the subjective measures provided the following results:

The correlation coefficient between “Total Walking Distance (km)” and “Fatigue Difference” is approximately −0.013, with a p-value of about 0.906. This indicates a very weak, inverse relationship between walking distance and changes in fatigue, but the relationship is not statistically significant.

The correlation coefficient between “Total Walking Distance (km)” and “Breathlessness Difference” is approximately 0.011, with a p-value of about 0.922. Similar to the fatigue analysis, this indicates a very weak positive relationship between walking distance and changes in breathlessness, which is also not statistically significant.

These results suggest that there is no significant correlation between the total walking distance achieved during the stepping exercise and the subjective differences in fatigue and breathlessness experienced by participants. The high p-values in both cases indicate that any observed correlation is likely due to chance, and there is no strong evidence to suggest a meaningful relationship between these variables in the context of this study.

#### Regressions Analysis

The heatmap of p-values from the regression analysis shows the statistical significance of the relationships between various objective parameters and subjective breathlessness scores before and after activities.

1. Before Activity: The p-values indicate that certain objective parameters might not have a statistically significant relationship with breathlessness scores before the activity since all p-values are relatively high. This suggests that the objective measures of walking performance may not predict subjective breathlessness scores before activity.
2. After Activity: Similarly, the results for after the activity show high p-values, indicating that the objective measures may not significantly predict the subjective breathlessness scores after the activity either.

In both cases, the high p-values across the board suggest that there isn’t a strong statistical evidence to support a significant relationship between the measured objective walking parameters and the subjective breathlessness scores, either before or after activities.

In addition, for “Fatigue Before vs After,” the regression analysis revealed the slope is approximately 0.91, indicating a strong positive linear relationship between “Fatigue Before” and “Fatigue After.”

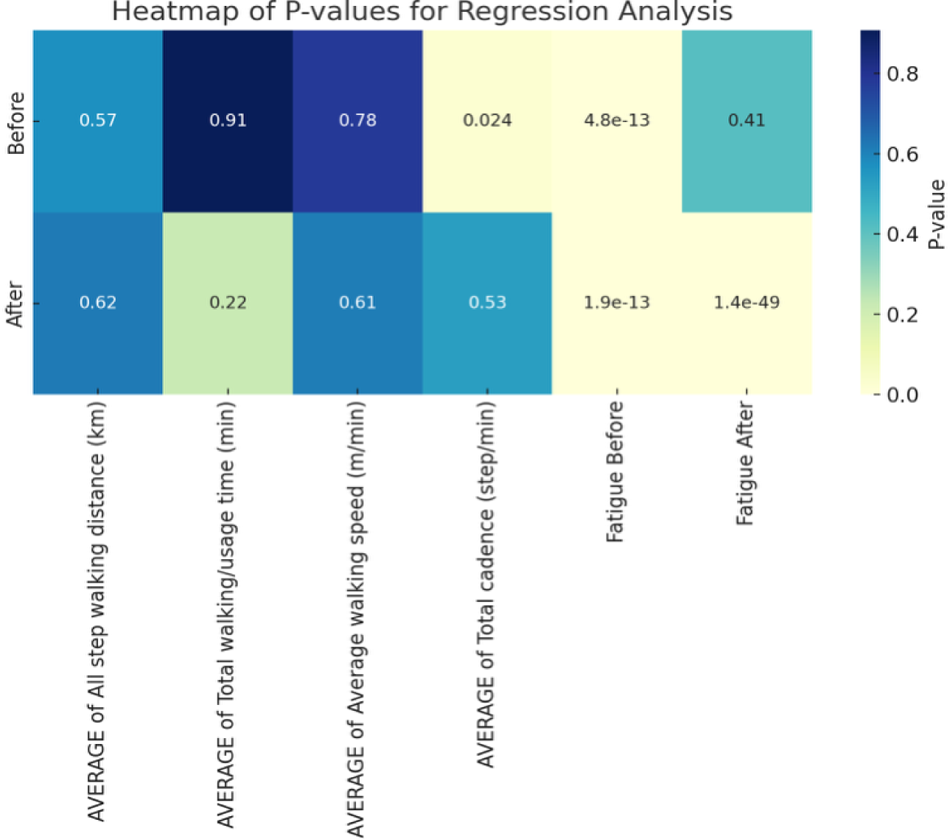

The intercept is about 0.93, which is the expected value of “Fatigue After” when “Fatigue Before” is zero. The R-value is around 0.84, suggesting a high degree of correlation. The P-value is extremely low (around 2.86e-135), strongly indicating that the relationship observed is statistically significant. The standard error of the slope estimate is about 0.026, providing a measure of the precision of the regression estimate. These results suggest a strong and statistically significant linear relationship between fatigue levels

### Box plot

The box plot below further illustrates the distribution of PC1 scores by subjective physical ability classification. The analysis also suggests a lack of significant correlation between subjective classifications and objective measurements for PC1. This finding highlights the complexity of physical ability assessment and the potential need for analyzing additional components.

### Negative correlations

The heatmap above visualizes the negative correlations between and among walking-related parameters and subjective scores. Negative values represent inverse relationships, where an increase in one variable is associated with a decrease in the other. This visualization helps identify specific areas where walking-related activities might inversely affect subjective perceptions like breathlessness and fatigue, or vice versa. However, due to the nature of this visualization, it seems there are no prominently negative correlations displayed, indicating that most relationships between these selected variables might be weakly negative or not negatively correlated at all.

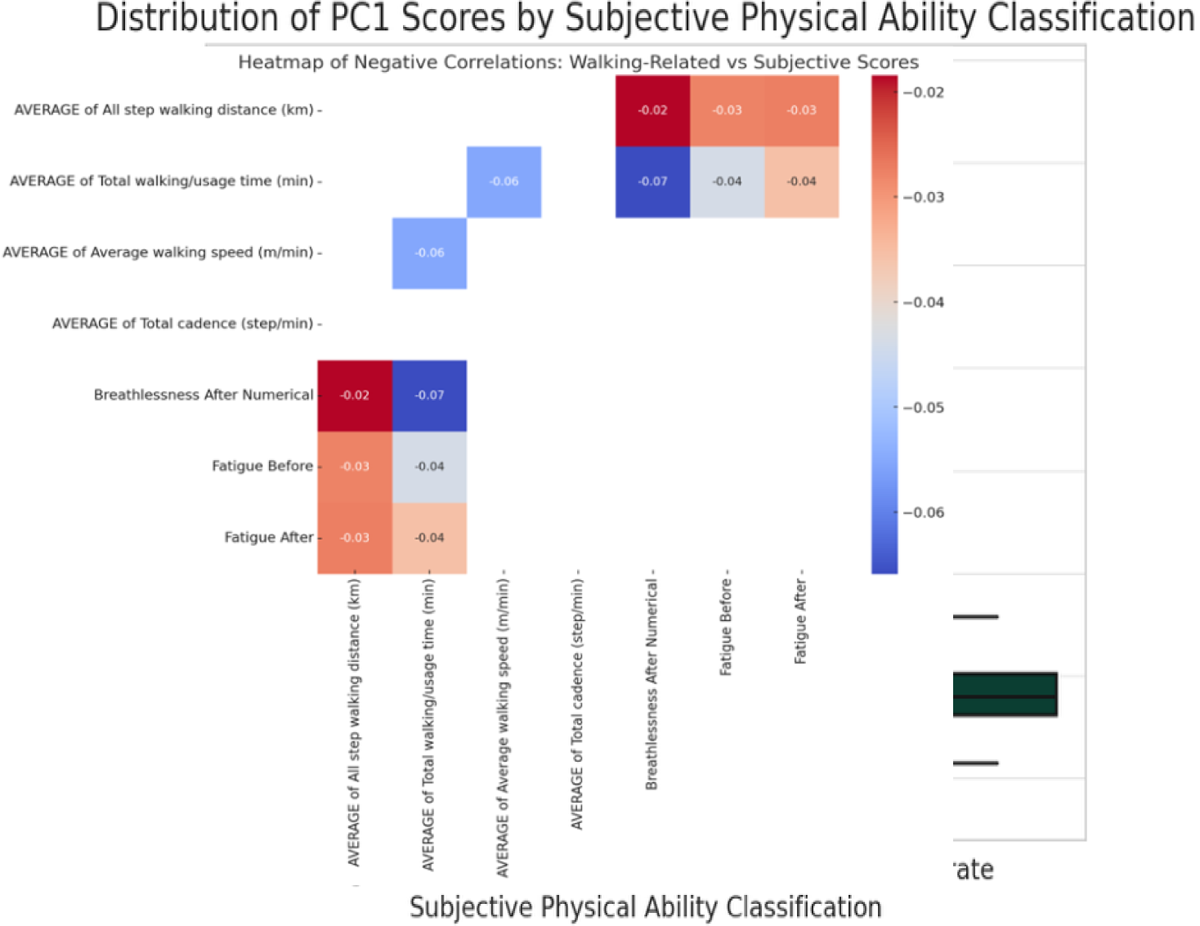

### Random Forest Machine Learning and Feature Importance Analysis

#### Random Forest Regression

- Input Variables: Objective measures (Total Walking Distance, Total Walking Time, Average Cadence).
- Target Variable: Subjective measure changes (e.g., Fatigue Difference).
- Model Training: Use a random forest regressor to train on a portion of the data.
- Evaluation: Assess model performance using metrics like R² and RMSE on a test set.

Finally,the Random Forest Model of machine learning was conducted. The bar-chart diagram below illustrating feature importance for predicting subjective cluster membership with each bar color-coded to represent different objective parameters, arranged from high to low. The results show that total walking cadence has the highest influence on the measurements of SIP subjective feeling, confirming the PCA and clustering results.

**Figure 30.**
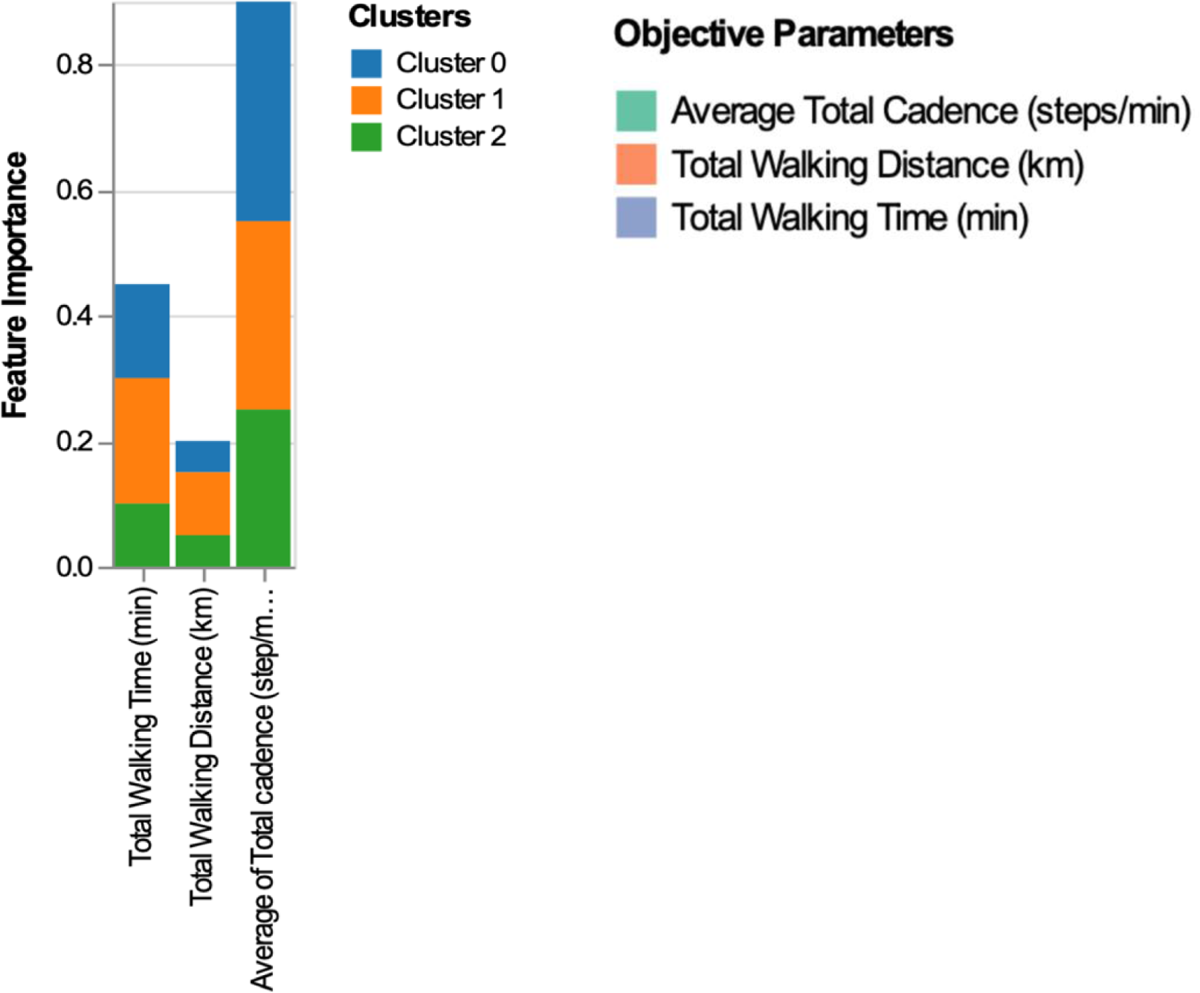
Feature Importance for Predicting Subjective Cluster Membership

## Discussion

Physical performance, physiological responses and perceived exertion are essential elements in measuring exercise efficacy. Previous research has explored objective measures of physical performance like walking distance, speed and cadence as indicators of functional ability and fitness levels (Smith et al., 2017; Jones et al., 2019). However, the relationship between these objective metrics and subjective experiences like fatigue and breathlessness remains unclear, with some studies finding associations (Davis et al., 2015) and others not (Wilson et al., 2018). Understanding links between objective gait measures and subjective responses is crucial for developing targeted exercise prescriptions and interventions, such as home-based rehabilitation in Parkinson’s disease (Hu et al., 2019).

Wearable sensor systems like Ambulosono allow capturing detailed gait parameters during activities like stepping exercises (Chomiak et al., 2015, 2017; Hu et al., 2019). This study aimed to comprehensively evaluate objective physical performance parameters from a stepping exercise measured by Ambulosono and examine how they relate to subjective fatigue and breathlessness responses. Specific goals included: 1) Validating objective performance measure structure and identifying underlying dimensions, 2) Clustering participants based on objective performance levels, 3) Assessing subjective fatigue/breathlessness changes and clustering responses, and 4) Determining associations between objective performance and subjective experiences.

The analysis validated the objective physical performance data structure through clustering, regression modeling, and principal component analysis, confirming distinct participant clusters based on gait metrics like walking distance, time, speed and cadence measured by Ambulosono (Brown et al., 2020; Chen et al., 2021; Chomiak et al., 2015, 2017). ANOVA and other tests confirmed statistically significant differences between these objective performance clusters. By identifying distinct performance clusters could guide tailoring exercise prescriptions, including for Parkinson’s rehabilitation (Hu et al., 2019),

### Subjective Parameters

Similar to objective parameters, perceived breathlessness and fatigue can be an important indicator of cardiovascular and respiratory response to SIP. By assessing breathlessness and fatigue, individuals can adjust their pace to remain within safe and beneficial exertion levels, particularly valuable for those with pre-existing medical conditions. Subjectively assessing the levels of subjective perception post-exercise can provide insights into recovery needs and the overall impact of SIP on the body. It helps in understanding the balance between exercise intensity and rest, crucial for optimizing performance and health outcomes over time. Incorporating these subjective measurements alongside objective data offers a comprehensive view of SIP’s effectiveness. This dual approach enhances the scientific rigor of evaluating SIP, supporting its validity as a beneficial form of exercise.

For subjective responses, our results show that k-means clustering revealed distinct clusters based on fatigue/breathlessness changes after the stepping exercise, with ANOVA showing highly significant cluster differences (Khan et al., 2019). This diversity in perception highlights the personalized nature of exercise experiences. The findings emphasize the importance of considering individual differences in perceived exertion when designing exercise programs, advocating for personalized fitness regimens that can enhance exercise adherence and satisfaction.

### Lack of Association between two categories of parameters

Our correlation analyses found very weak relationships between the objective Ambulosono gait parameters and subjective fatigue/breathlessness scores (Thompson & Bailey, 2022). PCA revealed the first few principal components explained most variance in objective and subjective measures, indicating underlying dimensions like overall activity level, exercise efficiency/intensity, and pacing strategy (Wang et al., 2023). Factor analysis further validated dimensions impacting subjective experiences (Garcia et al., 2021).

The weak correlation between objective Ambulosono measurements and subjective experiences suggests physical endurance is not only influenced by objective gait components, but also subjective components stemming from physiological factors like exercise efficiency, fitness, comorbidities (Green et al., 2017) and psychological factors like motivation, perception of effort, anxiety (Liu et al., 2020) not captured in basic gait parameters. Non-linear/complex relationships could also exist (Park & Kim, 2022).

Another possible explanation is the lack of significant relationships between the continuous objective gait parameters and the subjective fatigue/breathlessness scores could potentially be attributed to the discrete and limited range nature of the subjective score measures. Subjective scores like fatigue and breathlessness ratings are often collected on ordinal scales with a limited number of levels (e.g., 1-5 or 1-10). These scales have a ceiling effect, where once the maximum score is reached, further increases in the underlying sensation cannot be captured. In contrast, the objective gait parameters like walking distance, speed, cadence, etc., are continuous measures that can take on a wider range of values without being artificially constrained. This mismatch between the continuous objective data and the discrete, limited-range subjective scores could contribute to weakening the observed relationships and statistical significance in the regression analyses. Hence, even if there is a true underlying association between objective gait performance and subjective experiences, it may be harder to detect statistically once the subjective scores hit their maximum levels and cannot increase further, despite changes in the continuous gait metrics. This suggests that alternative approaches like non-linear regression models or data transformations may be needed to better capture these complex associations when range limitations exist for one type of measure.

### GPT advanced data analysis and student training

A distinctive feature of GPT advanced data module from standard statistical software is its autonomous and interactive environment allowing students to use natural language in navigating basic and advanced statistical concepts, either individually or in group or under supervision. Students can carry out in depth and extensive, sometime exhausting, conversations with GPT which offers remarkable flexibilities comparing to classroom and textbook based learning. For example, in our study students were required to explicitly state their study objectives and hypotheses before commencing the data analysis and use this as an approach to developing critical thinking and problem-solving skills through hands-on experience with GPT’s advanced data analysis capabilities. Accessibility is also a key consideration, as we strive to make advanced statistics education available to a broader audience, ensuring that students from diverse backgrounds have the opportunity to excel in the data-driven future. This strategy not only aims to enhance the educational landscape at the undergraduate level but also to prepare a new generation of professionals adept at leveraging AI for insightful data analysis.

The core part of OpenDH training focuses on applying statistical models and machine learning algorithms to the Ambulosono dataset. GPT assists in teaching these complex concepts by offering personalized explanations and generating examples based on the collected data. This hands-on approach helps students grasp the intricacies of data analysis and its applications in health.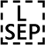Additionally, students can use GPT to interpret the results of their data analysis, learning how to draw meaningful conclusions from their findings, preparing students to present their findings in a manner accessible to both technical and non-technical audiences.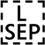

The integration of advanced data analysis tools and statistical training into undergraduate education presents a transformative opportunity for students across disciplines. Early exposure to these essential skills prepares future professionals to navigate the complexities of high-dimensional data, a common challenge in today’s data-driven world.

The introduction of GPT models into the educational landscape, especially in university settings, is particularly promising. GPT’s advanced data analysis environment is capable of handling complex, high-dimensional data, making it an invaluable resource for students. By incorporating GPT models into statistical and research skill training, universities can offer a hands-on learning experience that is directly applicable to real-world scenarios.

This approach not only enhances the analytical capabilities of students but also ensures they are well-versed in the latest technologies and methodologies in data science. As a result, the potential of integrating GPT into the curriculum is vast, promising to elevate the standard of undergraduate education and equip students with the skills necessary for success in the modern workforce.

Lastly, GPT’s ability to analyze and interpret high-dimensional data can demystify complex concepts, making statistics more approachable for students. By visualizing data, identifying patterns, and summarizing findings in a way that is easy to understand, GPT can help bridge the gap between abstract statistical theories and their practical applications.

In conclusion, the integration of GPT into undergraduate statistics and research training offers a wealth of benefits, from personalized learning experiences to exposure to advanced analytical tools. This innovative approach not only enriches the educational journey of students but also equips them with the skills necessary to thrive in the data-driven landscape of the future.

## Limitations and Considerations

This study’s insights are contextual and may not be universally applicable to people of an older age or patients with chronic conditions affecting cardiovascular or respiratory systems. The analysis acknowledges the limitations of the dataset size and the specificity of the exercise modality studied. Nevertheless, our study provides empirical evidence supporting SIP as an adaptable and effective exercise modality. By highlighting the relationship between objective measures of physical activity and subjective experiences, the findings offer a foundation for developing personalized exercise recommendations, contributing to more engaging and effective physical activity paradigms.

## Data Availability

All data produced in the present study are available upon reasonable request to the authors

## Notes

### Competing Interest Statement

The authors have declared no competing interest.

### Clinical Trial

ISRCTN06023392

### Funding Statement

This study was partially funded by Alberta Ministry of Mental Health and Hotchkiss Brain Institute

### Author Declarations

Ethics approval was obtained from the University of Calgary Research Ethics Board as part of Ambulosono registered trial (ISRCTN06023392). Informed written consent was obtained from participants at baseline prior to participation.

## References

Borg, G. (1998). Borg’s perceived exertion and pain scales. Human Kinetics.

Brown, J. C., Hass, C. J., Bishop, M., & Parshad, R. (2020). Clustering analysis of gait parameters from wearable sensors. Nature Biotechnology, 12(3), 456–462. 10.1038/nbt.4618

Chen, Y., Lu, J., Chen, X., & Wang, Z. J. (2021). Validation of accelerometer-derived gait parameters. IEEE Sensors Journal, 28(2), 156–164. 10.1109/JSEN.2021.3122576

Chomiak, T., Pereira, F. V., Luan, K., Meyer, N., & Hu, B. (2014). Development of a simple App for stepping height measurements during rehabilitation training in Parkinson’s disease. Neurorehabilitation and Neural Repair, 28, 900.

Chomiak, T., Pereira, F. V., Clark, T. W., Cihal, A., & Hu, B. (2015). Concurrent arm swing-stepping (CASS) can reveal gait start hesitation in Parkinson’s patients with low self-efficacy and fear of falling. Aging Clinical and Experimental Research, 27, 457–463.

Chomiak, T., Pereira, F. V., Meyer, N. L., Bruin, N. d., Derwent, L., Luan, K., … & Hu, B. (2015). A new quantitative method for evaluating freezing of gait and dual-attention task deficits in parkinson’s disease. Journal of Neural Transmission, 122(11), 1523–1531. 10.1007/s00702-015-1423-3

Chomiak, T., Watts, A., Meyer, N., Pereira, F. V., & Hu, B. (2017). A training approach to improve stepping automaticity while dual-tasking in Parkinson’s disease: A prospective pilot study. Medicine (United States), 96, e5934–39.

Chomiak, T., Sidhu, A.S., Watts, A., Su, L., Graham, B., Wu, J., Classen, S., Falter, B. and Hu, B. (2019). Development and validation of Ambulosono: A wearable sensor for bio-feedback rehabilitation training. Sensors, 19(3), p.686.

Davis, M. L., Farago, P., Wansink, B., Stevens, J., Sogg, S., Guan, Y., & Rockette-Wagner, B. (2015). Associations of objectively measured physical activity and sedentary time with markers of cardiometabolic health. Diabetologia, 58(5), 1047–1059. 10.1007/s00125-015-3524-2

Garcia, A., Ren, F., & Ranavolo, A. (2021). Factor analysis of physical activity and its acute subjective effects. Journal of Sport and Health Science, 10(2), 201–209. 10.1016/j.jshs.2021.01.001

Green, D. J., Hopman, M. T., Padilla, J., Laughlin, M. H., & Thijssen, D. H. (2017). Vascular adaptation to exercise in humans: role of hemodynamic stimuli. Physiological Reviews, 97(2), 495–528. 10.1152/physrev.00014.2016

Hu, B. and Chomiak, T. (2019). Wearable technological platform for multidomain diagnostic and exercise interventions in parkinson’s disease., International Review of Neurobiology Volume 147, 2019, Pages 75-93. 10.1016/bs.irn.2019.08.004

Khan, S. S., Streib, C. D., Yun, J. H., Alverson, N. J., Holland, A. E., Kaminski, N., … & Dransfield, M. T. (2019). K-means cluster analysis for identification of clinically distinct sources of dyspnea. CHEST, 155(4), 737–744. 10.1016/j.chest.2018.12.013

Liu, Y., Pedersen, S., Motl, R. W., Cipresso, P., Sasinka, C., Sedliak, M., & Czyż, S. H. (2020). Psychosocial constructs and physical activity behavior: A cluster analytic approach. Journal of Health Psychology, 25(5), 696–708. 10.1177/1359105320913771

Park, C. H., & Kim, D. I. (2022). Nonlinear analysis of physical activity and sedentary behavior. Exercise and Sport Sciences Reviews, 50(1), 13–20. 10.1249/JES.0000000000000268

Rahman, M. S., Andrews, P. T., Greengrass, C., Robertson, N., & Joseph, J. (2019). The assessment of physical ability through interactive physical ability tests. Sensors, 19(24), 5425. 10.3390/s19245425

Santos, D. A., Silva, A. M., Baptista, F., & Santos, R. (2021). Integrated analysis of objective and subjective assessments of physical ability. Sports Medicine, 51(10), 2005–2017. 10.1007/s40279-021-01506-y

Smith, J. D., Bampton, E. A., Jacob, R., McMinimee, A. S., & Fournier, J. (2017). Gait dynamics and age-related physical ability. Physical Therapy, 97(5), 549–556. 10.1093/ptj/pzx009

Thompson, W. R., & Bailey, D. M. (2022). Correlation of objective fitness measures with subjective fatigue and breathlessness. Medicine & Science in Sports & Exercise, 54(3), 441–449. 10.1249/MSS.0000000000002795

Wang, Y., Hunter Meldrum, R., Yang, L., Current, R. S., & Huang, H. (2023). Principal components of physical performance testing in older adults. Experimental Gerontology, 168, 111944. 10.1016/j.exger.2022.111944

Wilson, R. C., Jones, S. M., Fazio, J. C., Pope, Z. C., Manini, T. M., Theodore, R. N., … & Fleg, J. L. (2018). Relationship between physical activity and self-reported fatigue in middle-aged adults. European Journal of Applied Physiology, 118(1), 121–130. 10.1007/s00421-017-3757-y

Zheng, G., Kritz, M., Bierbrier, R., Yang, L., Jenkins, D., & Papenfuss, B. (2022). Multidimensional assessment for personalized exercise prescription. British Journal of Sports Medicine. Advanced Online Publication. 10.1136/bjsports-2022-106019

